# Proteomic and Genetic predictors and risk scores of cardiovascular diseases in persons living with HIV

**DOI:** 10.1101/2025.05.08.25327219

**Authors:** Tanvi Mehta, Lillian Haine, Jason Baker, Cavan Reilly, Daniel Duprez, Shweta Mistry, Brian Steffen, Mamta K. Jain, Alejandro Arenas-Pinto, Mark Polizzotto, Therese Staub, Sandra E. Safo, INSIGHT ESPRIT, FIRST, SMART and START study groups

**Author notes:** Correspondence: Sandra E. Safo, Division of Biostatistics and Health Data Science, School of Public Health, 2221 University Avenue SE, Suite 200, University of Minnesota, Minneapolis, MN 55414 USA.

## Abstract

**Background:** Cardiovascular diseases (CVD) prediction models for persons living with HIV (PLWH) depend on traditional CVD risk factors, but these underestimate true risk. We aimed to identify proteins and genetic variants and create proteo-genomic risk scores for CVD in PLWH.

**Methods:** We analyzed genetic and protein data from participants involved in trials for PLWH. We used state-of-the-art statistical methods for data integration, identified correlated signatures, and developed a protein score (PS) and a genetic score (GS) to predict CVD. We conducted functional enrichment analysis to explore biological functions of signatures identified in relation to CVD.

**Results:** A panel of 14 proteins and a set of 15 genetic variants were found to be better at distinguishing between CVD cases and controls than individual proteins or genetic variants. The PS or GS was each independently associated with a higher risk of CVD. Combining CVD-, HIV-related factors, genetics, and protein scores resulted in the most powerful discrimination. Functional enrichment analysis showed an upregulation of the cytokine tumor necrosis factor (TNF) and strong enrichment for inflammation related pathways such as the pathogen induced cytokine storm.

**Conclusions:** A panel of protein biomarkers, some new (IGFBP7, HGF) and some previously known in PLWH (CLEC6A), could help identify PLWH at higher risk of developing CVD. If confirmed, these scores could be used with CVD and HIV-related factors to identify PLWH at risk for CVD who would benefit from proactive risk reduction strategies.

## Introduction

It is well known that people living with HIV (PLWH) are at an elevated risk for cardiovascular diseases (CVDs) when compared to those without HIV^1,2^. In a recent systematic review investigating CVD in PLWH, it was found that PLWH are twice as likely to develop CVD compared to those without HIV^3^. PLWH are still at an elevated risk for CVDs when compared to HIV negative controls, even with viral suppression and effective combination antiretroviral therapies^4^. This increased risk has been consistent across multiple studies and CVD related outcomes, including myocardial infarction^5,6^ and heart failure^7^. With effective combination antiretroviral therapies (cARTs) HIV is now a chronic, manageable disease, with the primary driver of morbidity and mortality in PLWH shifting from opportunistic infections to chronic, age-related comorbidities, such as CVDs^4^. This has resulted in a steadily increasing global burden of HIV-associated CVDs, which has tripled over the last two decades^3^. As such, characterizing the increased risk of CVD in PLWH is an increasingly important question.

Current clinical guidelines for CVD management in PLWH use traditional risk factors and CVD risk prediction scores that have been developed in the general population^4^. This generally involves modification of traditional CVD lifestyle risk factors such as lifestyle modifications via diet, exercise, and limiting alcohol and tobacco use^4^. With HIV-specific CVD management the use of commonly prescribed statins and blood pressure lowering medication is complicated due to ART and statin drug-drug interactions^4^. Additionally, CVD risk prediction using traditional CVD factors underestimates the risk in PLWH and so are less effective at characterizing CVD risk in PLWH^8^. As such, finding new ways to better characterize CVD risk in PLWH is critical.

With technological and statistical advancements, the collection of multi-omics data such as genomics, proteomics, and metabolomics data is becoming more widely accessible. Due to the complex nature of CVDs polygenic risk scores have recently been proposed as a way to improve risk stratification for CVD^9,10^. Polygenic risk scores are able to more effectively characterize CVD risk in the general population, however the utility of polygenic risk scores is questioned in the general clinical setting as they often require millions of SNPs to be measured, which is often infeasible in the clinical setting. However, developing a more focused set of SNPs to use might more readily serve the clinical setting. Similarly, proteomic data and the development of proteomic risk scores has recently been proposed as an alternative approach to improve CVD risk stratification without necessitating measurement of most, if not all, of the genome. Thus, using approaches that leverage both more targeted genetic data and proteomic data might provide a clinically useful approach that allows for improved CVD risk stratification.

Here, we posited that leveraging information from both genomic and proteomic data to create a CVD proteo-genomic risk score, using a statistical method such as the sparse integrative discriminant analysis method (SIDA)^11^, might be more able to distinguish cases and controls in PLWH than a traditional risk score that uses only proteomic or genomic data. We also hypothesized that the scores would predict CVD better than the individual components used to develop the scores. We use focused genetic data by pulling only SNPs that are on genes that have been found to be related to inflammation; this allows us to pre-specify and pull a subset of SNPs, which will be more clinically relevant and allow for a more feasible computation time for SIDA. With the proposed approach, we believe that we will be able to better understand and characterize CVD risk in PLWH than using only genetic or proteomic data alone.

## Methods

### Population and Outcome Variable

Participants were enrolled in one of four trials conducted by the Terry Beirn Community Programs for Clinical Research on AIDS (CPCRA) and the International Network for Strategic Initiatives in Global HIV Trials (INSIGHT): FIRST^12^, ESPRIT^13^, SMART^14^, or START^15^. Enrollment in these trials occurred from 1999 to 2013. The results and population under study for each of these trials have been published previously^12–15^. CVD case status was defined as non-fatal stroke, non-fatal myocardial infarction, coronary revascularization, or death from CVD, including unwitnessed deaths at any time during follow-up. Controls were matched based on age (+/-5 years) at baseline, treatment arm of study, and randomization date (+/-90 days). All cases and controls consented to the collection of DNA and have genomics data and to the collection and storage of plasma samples at study entry that were used to measure protein biomarkers.

### Genomics Data, Pathways and Quality Control

The genotyping process has been published previously^2^. Briefly, participants were genotyped using a custom Affymetrix Axiom SNP array with 770,□558 probe sets enriched for markers related to immune function; we kept SNPs for our analysis that were directly measured and passed quality control, which left us starting with 655,641 SNPs. SNP quality control is depicted in the flow chart given in Figure S1 in the supplementary material. Briefly, starting with 655,451 genetic variants, we used dbSNP to identify variants and the base positions (starting locus and stopping locus) for genes associated with the inflammation pathway (61,694 variants). We used the base pair locations for these genes to identify SNPs associated with the inflammation pathway; SNPs were only included if they were in the base pair location interval specified by dbSNP. We removed variants with minor allele frequency (MAF) < 0.05, not in Hardy-Weinberg Equilibrium (p-value < 10E-06), and any variants with missingness. This resulted in 9,166 variants for analyses.

### Proteomics Data from Olink

We used a baseline plasma specimen from individuals who consented to genomics to measure protein biomarkers from 5 Olink multiplex panels (Cardiovascular II, Cardiovascular III, Immune Response, Inflammation, and Cardiometabolic). Each panel has 92 target proteins. Some proteins appear in multiple panels. Sample analysis and detection of proteins from Olink is performed using proximity extension analysis (PEA) with the target protein detected through high-throughput real-time polymerase chain reaction (PCR)^16^. The PEA assay by Olink has been used in the literature for identifying risk factors for CVD.

### Traditional Risk Factors

CVD specific risk factors considered were: sex at birth; baseline age; self-reported race categorized as either Black, Hispanic, Asian, White, or Other for SMART, START, and FIRST, with ESPRIT as either Black, Asian, White, or Other; history of CVD at baseline, defined as prior coronary artery disease (CAD) requiring treatment, prior MI, prior stroke, or prior CAD requiring surgery; taking lipid lowering medication at baseline; taking blood pressure lowering medication at baseline; diabetes, defined as a diagnosis of diabetes requiring drug treatment. HIV specific baseline factors that were considered were: ART use, CD4+ values, and HIV RNA values.

## Statistical Analysis

### Integrative Analysis of Genetics and Proteomics Data

We considered a multivariate approach to investigate the associations between the genetics and proteomics data, and to determine genetic variants and proteins that are correlated and discriminate between CVD cases and controls. Proteins that appeared on multiple panels were all considered in the integrative analysis. We used the sparse integrative analysis (SIDA) approach^11^ for this purpose, and coupled SIDA with bootstrap resampling for statistical rigor. We embedded a univariate logistic regression filtering step into the integration step to ensure that variables that went into SIDA discriminated between CVD cases and controls. Refer to supplementary material for more details. Proteins and genes passing the univariate filtering step and were frequently selected by SIDA to discriminate between cases and controls were chosen as candidate variables for downstream analyses.

### Pathway Enrichment Analysis

We used Ingenuity Pathway Analysis [IPA] (QIAGEN) to perform functional enrichment analysis of proteins detected to be associated with genetic variants in the multivariate integrative analysis approach based on SIDA. We used the core analysis functionality in IPA to predict involvement of signaling and metabolic pathways and to predict upstream molecules regulating the proteins we identify^17^.

### Development of Protein and Genetic Risk Scores

We developed molecular scores (i.e. protein ang genetic scores) to assess the contribution of the identified biomarkers in a single combined measure. For each bootstrap training dataset and each biomarker, we used a logistic regression model to obtain the log odds ratio of the biomarker in discriminating CVD case status. A weighted mean was then used to obtain an overall log-odds ratio for each biomarker. We used the full dataset and the weighted mean for each protein to derive the protein score. We also developed an ancestry adjusted SNP score. To do this we obtained the residuals from a multiple linear regression model using the top four principal components (PCs) from the principal components analysis to predict the unadjusted SNP score. The PCs were calculated from genome-wide SNPs for our case-control samples. The distribution of the scores is described using median and interquartile range (IQR) (Table 1) and compared across different demographic variables (Figures S4-S6).

**Table 1:** Baseline Characteristics for CVD Cases and Controls.

| **Characteristic** | **Overall,**<br>**N = 360<sup>1</sup>** | **Case,**<br>**N = 122<sup>1</sup>** | **Control,**<br>**N = 238<sup>1</sup>** | **p-value<sup>2</sup>** |
| --- | --- | --- | --- | --- |
| **Gender** |  |  |  | 0.08 |
| Female | 43 (12%) | 9 (7%) | 34 (14%) |  |
| Male | 317 (88%) | 113 (93%) | 204 (86%) |  |
| **Study** |  |  |  | >0.9 |
| ESPRIT | 215 (60%) | 74 (61%) | 141 (59%) |  |
| FIRST | 10 (2.8%) | 3 (3%) | 7 (2.9%) |  |
| SMART | 45 (12.5%) | 15 (12%) | 30 (13%) |  |
| START | 90 (25%) | 30 (25%) | 60 (25%) |  |
| **CVD at Baseline** | 18 (5%) | 15 (12%) | 3 (1.3%) | <0.001 |
| **Age (YRS)** | 48 (42, 54) | 49 (41, 55) | 47 (42, 54) | 0.41 |
| **Black Race** |  |  |  | >0.9 |
| Black | 56 (16%) | 19 (16%) | 37 (16%) |  |
| Non Black | 304 (84%) | 103 (84%) | 201 (84%) |  |
| **Race/Ethnicity** |  |  |  | 0.45 |
| Asian | 1 (0.3%) | 1 (0.8%) | 0 (0%) |  |
| Black | 56 (16%) | 19 (16%) | 37 (16%) |  |
| Hispanic | 16 (4.4%) | 5 (4.1%) | 11 (4.6%) |  |
| Other | 8 (2.2%) | 1 (0.8%) | 7 (2.9%) |  |
| White | 279 (78%) | 96 (79%) | 186 (77%) |  |
| **Diabetes**<br>**Diagnosis at**<br>**Baseline** |  |  |  | 0.32 |
| No | 346 (92.8%) | 114 (90.5%) | 232 (93.9%) |  |
| Yes | 27 (7.5%) | 12 (9.8%) | 15 (6.3%) |  |
| **CD4 at Baseline** | 546 (406, 668) | 551 (411, 631) | 545 (404, 683) | 0.53 |
| **Lipid Lowering Treatment at Baseline** |  |  |  | 0.10 |
| No | 290 (81%) | 92 (75%) | 198 (83%) |  |
| Yes | 70 (19%) | 30 (25%) | 40 (17%) |  |
| **Blood Pressure Treatment at Baseline** |  |  |  | 0.006 |
| No | 295 (82%) | 90 (74%) | 205 (86%) |  |
| Yes | 65 (18%) | 32 (26%) | 33 (14%) |  |
| **ART at Baseline** |  |  |  | 0.67 |
| No | 104 (28.9%) | 33 (27.0%) | 71 (29.8%) |  |
| Yes | 256 (71.1%) | 89 (73.0%) | 167 (70.2%) |  |
| **Proteomic Risk Score** | 33.56 (31.84, 35.24) | 34.72 (33.37, 36.40) | 33.03 (31.52, 34.55) | <0.001 |
| **SNP Risk Score** | 0.46 (-0.64, 1.58) | 1.53 (0.62, 2.69) | -0.09 (-0.81, 0.78) | <0.001 |
| *I* n (%); Median (IQR)<br>2 Pearson's Chi-squared test; Wilcoxon rank sum test; Fisher's exact test |  |  |  |  |

### Model Development and Assessment

We used multivariable logistic regression models to investigate the model prediction performance of the following models: (i) baseline, (ii) baseline + individual proteins, (iii) baseline + individual SNPs, (iv) baseline + protein score, (v) baseline + genetic score, and (vi) baseline + protein score + genetic score. The baseline model included both HIV related factors, the CD4 value at baseline, and CVD related factors, gender, age, diabetes status, an indicator for Black race, prior history of CVD, and lipid and BP lowering medication variables all measured at baseline. The genetic model adjusted for ancestry using the first 4 PCs. We demonstrated the performance of the model in terms of discrimination, predictive ability, and calibration. For discrimination, we used the area under the receiving operating characteristic curves (AUC) and the continuous or categorical free net reclassification improvement (NRI)^19–21^ to assess whether adding the score (protein score, genetic score, or both) to the baseline model improved discrimination more than the baseline model only or adding each individual biomarker included in the scores to the baseline model. For predictive ability, we considered the effect sizes (odds ratios) of the scores and whether they were statistically significant in discriminating CVD cases and controls. The Hosmer-Lemeshow goodness of fit test was used for calibration. We performed all analyses using R software version 4.2.1 (The R Foundation). Statistical significance was ascertained at a 2-sided significance level of 0.05 and since these are exploratory analyses, we did not perform any adjustment for multiplicity.

## Results

### Study Population

There were 360 individuals included in the analyses, 122 with CVD (cases) and 238 without CVD (controls). Twelve percent of participants are female, 0.3% are Asian, 16% are Black, 4.4% are Hispanic, and 78% are White. The median age at baseline is 48 (IQR: 42-54), and the median CD4 cell count at baseline is 546 (IQR: 406-668). At baseline, 5% of participants had a history of a CVD event, 7.5% of participants had a diabetes diagnosis, 19% of participants were on a lipid lowering treatment, and 18% of participants were on a blood pressure treatment. The median proteomic risk score is 33.56 (IQR: 31.84-35.24), and the median SNP risk score is 0.46 (IQR: −0.64-1.58). These characteristics are presented overall and by case/control status in Table 1.

### Multivariate Proteo-genomic Integrative Analysis to Determine Correlated Proteins and Genetic Variants Discriminating CVD Cases and Controls

Multivariate integrative analysis using SIDA identified 16 (14 unique) proteins, including IL6 protein from 3 Olink panels, and 15 genetic variants as being able to discriminate between CVD cases and controls. All IL6 from the 3 Olink panels were selected by SIDA but we only kept the IL6 with maximum expression in subsequent analyses. The 14 unique proteins were: IL-6, CCL11, CLEC6A, HGF, FGF19, ADAMTS13, IL1RL2, GT, CCL18, PLA2G7, LTBR, UPAR, SCGB3A2, IGFBP7. Six of these proteins (IL6, CCL11, SCGB3A2, GT, PLA2G7, HGF) were identified in our previous work that used proteomics data only^24^. We provide an overview of the literature for each protein in the supplementary material.

### Pathway Analysis of Molecules Identified in Proteo-genomic Integrative Analysis

The 14 unique proteins identified by SIDA were input into String^18^ for protein-protein interactions and IPA for functional enrichment analyses to predict involvement of signaling and metabolic pathways. We included the log-odds ratios for each molecule from univariate logistic regression (Table S1). Figure 1 suggests that the proteins identified by the proteo-genomic analysis are biologically related as a group, with a protein-protein enrichment p-value of 5.46E-0.9. In Figure 2 we show IPA analyses for the molecules that are shared among the canonical pathways, with a focus on the top 10 significantly enriched pathways having at least two shared molecules. Some pathways included the pathogen induced cytokine storm signaling pathway, tumor microenvironment pathway, the granulocyte adhesion and diapedesis pathway and the atherosclerosis signaling pathway. The three proteins found in the atherosclerosis signaling pathway (CCL11, IL6, PLA2G7) were each significantly associated with an increased CVD risk in a univariate analysis (p-value <0.05, OR range 1.3-1.5) [Table S1]. Regarding upstream regulators, tumor necrosis factor (TNF), lipopolysaccharide (LPS) and tetradecanoylphorbol acetate were each predicted to be activated. In particular, the cytokine TNF was predicted to be activated with a z-score of 2.077 and overlap p-value 5.65E-06. Six (HGF, LTBR, IL6, UPAR, CCL11, GT) out of 8 genes known in the literature to be upregulated by TNF were also upregulated in our dataset, which is consistent with activation of TNF.

**Figure 1.**
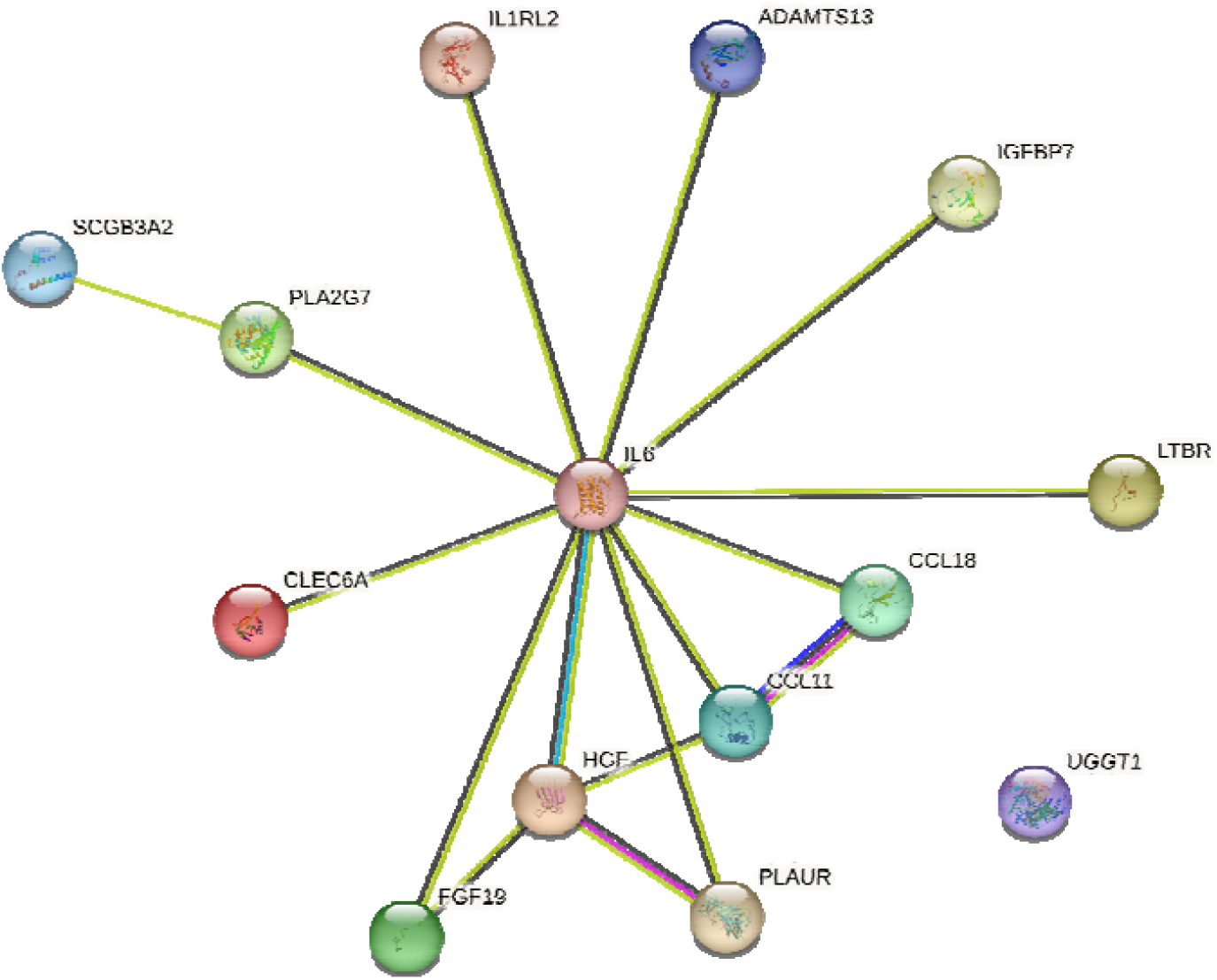
Network of fourteen proteins from proteo-genomics analysis. Source: https://string-db.org/. Nodes: UGGT1 is GT, PLAUR is UPAR. Meaning of lines: blue: from curated databases, pink: experimentally determined, green: gene neighborhood, red: gene fusions, blue: gene co-occurrence, light green: text mining, black: co-expression.

**Figure 2.**
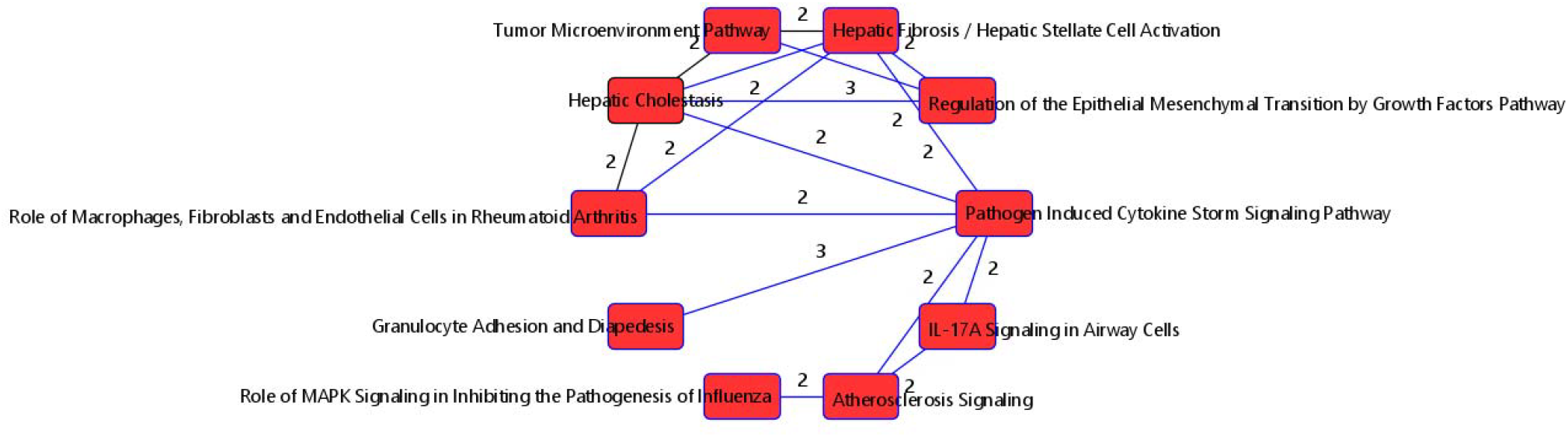
Top 10 Overlapping Canonical Pathways from IPA for Proteins Discriminating Between CVD Cases and Controls in Multivariate Integrative Analyses. Edge numbers indicate the number of molecules that are shared between the pathways connected by the edge.

## Model Development and Assessment

### Comparison of baseline and protein models

An increase in the protein score by one standard deviation is associated with an odds ratio for CVD of 2.36 (CI: 1.78 to 3.19, p-value < 0.05), adjusting for baseline variables [Table S2]. When standardized and added to the baseline model individually, all 14 proteins have ORs that are lower than 2.36 (the OR for the protein score in the baseline + protein score model), and 13 out of 14 proteins (IL-6, CCL11, CLEC6A, HGF, FGF19, ADAMTS13, GT, CCL18, PLA2G7, LTBR, UPAR, SCGB3A2, IGFBP7) are statistically significant (p-value < 0.001) [Table S3]. Because the proteins and the protein score have been standardized, we are able to directly compare the ORs of the individual proteins from the individual protein models to that of the protein score (Table S3) in the protein score model. The baseline model has an AUC of 0.61 (CI: 0.55 to 0.67) and the baseline + protein score model has an AUC of 0.74 (CI: 0.69 to 0.80), which is a 21.3% improvement in AUC or a 54.2% improvement in prediction ability. All individual protein models have lower AUCs and NRIs than that of the baseline + protein score model (Tables 4 and S4). The overall net reclassification improvement (NRI) is 0.65 (CI: 0.44 to 0.86); cases have an NRI of 0.31 (CI: 0.13 to 0.48) and controls have an NRI of 0.35 (CI: 0.22 to 0.47). This suggests that addition of the protein score to the baseline model increases true positive rate (ability to correctly predict a case) by 31% and reduces false positive rate by 35%. When analyses were restricted to individuals on ART at baseline, the protein score was again statistically significant (OR: 2.27, CI: 1.65 to 3.20, p-value < 0.001) [Table S5]. The Hosmer-Lemeshow goodness of fit test suggests that the protein score model fits the data well (p-value > 0.05). There were no statistically significant interactions between the protein score and cd4 count at baseline (treated as continuous or categorized into high [> 500 cell/mm^3^] vs low [<= 500 cell/mm^3^], p-value > 0.05).

**Table 4:** Incremental Contribution of Protein and Genetic Scores to CVD Risk When Added to Baseline Model (n=360). LCI: lower confidence interval. UCI: upper confidence interval.

| Model | AUC | LCI | UCI | NRI Cases | NRI Controls | Overall NRI |
| --- | --- | --- | --- | --- | --- | --- |
| Baseline Model | 0.612 | 0.550 | 0.675 |  |  |  |
| Baseline + Protein Score | 0.742 | 0.687 | 0.797 | 0.305 | 0.345 | 0.65 |
| Baseline + SNP Score + 4 PCs | 0.829 | 0.784 | 0.874 | 0.492 | 0.467 | 0.959 |
| Baseline + Protein Score + SNP Score + 4 PCs | 0.858 | 0.819 | 0.897 | 0.525 | 0.546 | 1.071 |

### Comparison of baseline and genetic models

A similar multivariable logistic regression model was fitted using the standardized SNP score to predict CVD case status, adjusting for the first 4 PCs in addition to the factors listed above [Table S6]. An increase in the SNP score by one standard deviation is associated with an odds ratio for CVD of 4.59 (CI: 3.21 to 6.80, p-value < 0.001). All 15 SNPs individually have ORs lower than 4.59, and 14 out of 15 SNPs (rs11895665, rs2240688, rs4696483, rs34308112, rs10456432, rs3808528, rs17252559, rs3816208, rs9410490, rs115708227, rs17053844, rs80067004, rs73320494, rs12602462) are statistically significant (p-value < 0.05) (Table S7). The baseline + 4 PCs + SNP score model has an AUC of 0.83 (CI: 0.78 to 0.87), which is a 36.1% improvement in AUC or a 66.7% improvement in prediction ability compared to the baseline model. All individual SNP models have lower AUCs and NRIs than that of the baseline + 4 PCs + SNP score model (Table S4). The overall NRI is 0.96 (CI: 0.76 to 1.15); cases have an NRI of 0.49 (CI: 0.33 to 0.65) and controls have an NRI of 0.47 (CI: 0.35 to 0.58). When analyses were restricted to individuals on ART at baseline, the genetic score was again statistically significant (OR: 3.82, CI: 2.63, 5.81, p-value < 0.001) [Table S8]. The Hosmer-Lemeshow goodness of fit test suggests that the genetic score model fits the data well (p-value > 0.05). There were no statistically significant interactions between the genetic score and cd4 count at baseline (treated as continuous or categorized into high [> 500 cell/mm^3^] vs low [<= 500 cell/mm^3^], p-value > 0.05).

### Comparison of baseline and genetic + protein models

A comparable multivariable logistic regression model was fitted using both the standardized protein score and standardized SNP score to predict CVD case status, adjusting for 4 PCs in addition to the baseline factors. Holding the SNP score and all other variables constant, an increase in the protein score by one standard deviation is associated with an odds ratio for CVD of 2.28 (CI: 1.64 to 3.25) [Figure 3] and, holding the protein score and all other variables constant, an increase in the SNP score by one standard deviation is associated with an odds ratio for CVD of 4.73 (CI: 3.25 to 7.25) [Figure 3]. The baseline + protein score + 4 PCs + SNP score model has an AUC of 0.86 (CI: 0.82 to 0.90) [Table 4], which is a 41.0% improvement in AUC or a 69.4% improvement in prediction ability when compared to the baseline model. The overall NRI is 1.07 (CI: 0.88 to 1.26); cases have an NRI of 0.53 (CI: 0.37 to 0.68) and controls have an NRI of 0.55 (CI: 0.44 to 0.65). When analyses were restricted to individuals on ART at baseline, the genetic score and protein score were each independently statistically significant (Table S9). The Hosmer-Lemeshow goodness of fit test suggests that the protein and genetic score model fits the data well (p-value > 0.05).

**Figure 3.**
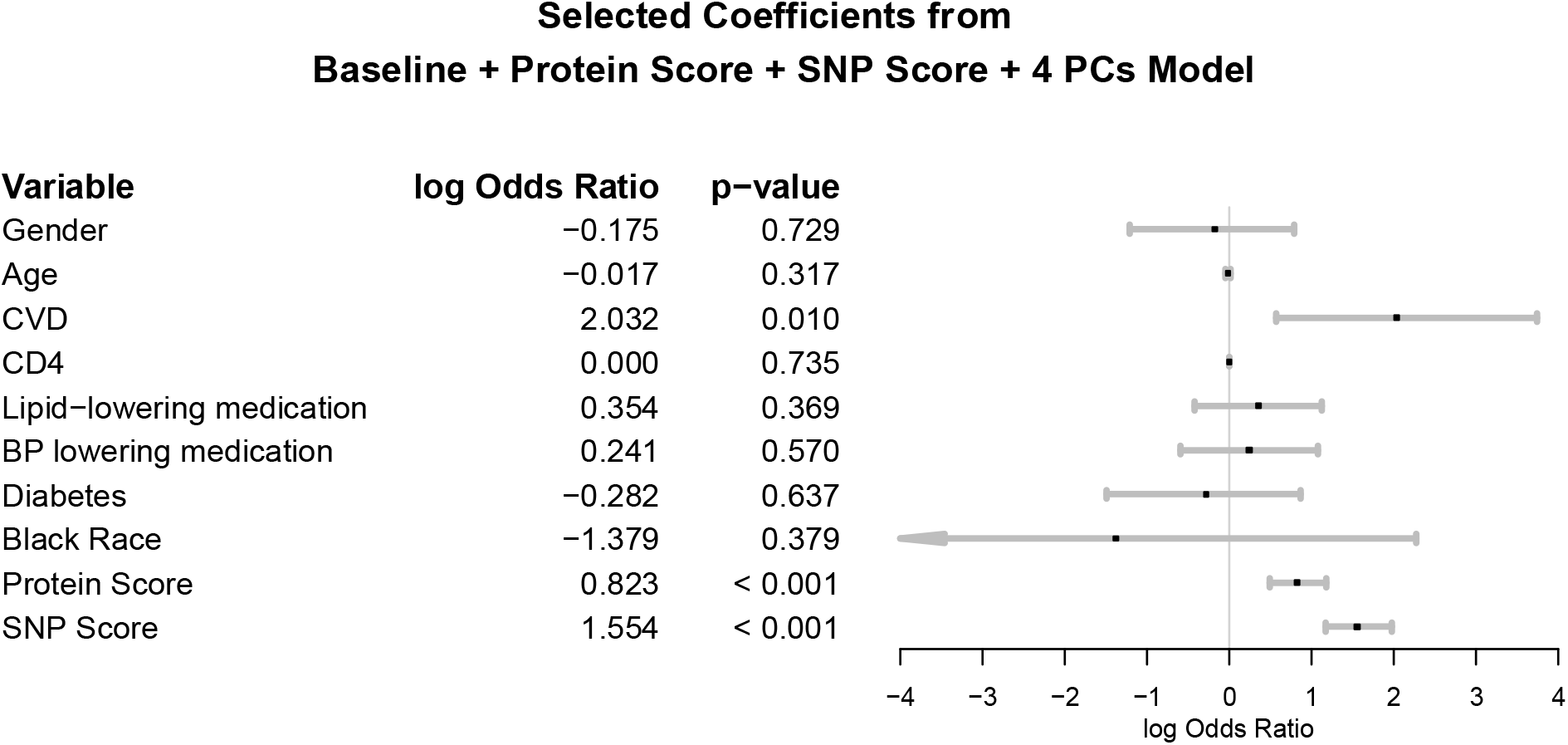
Forest plot for baseline + protein score+ SNP score, adjusted for first four principal components.

**CD4: CD4+ count at baseline Race: race was self-reported race**.

**CVD is measured at baseline: history of cardiovascular disease at baseline defined as prior coronary artery disease (CAD) requiring treatment, prior MI, prior stroke, or prior CAD requiring surgery**

Baseline is a model with the following variables: CD4, sex, age, diabetes status at baseline, prior history of cardiovascular disease, lipid□lowering medication, blood pressure□lowering medication, and a black race indicator. NRI (cases and controls are proportions, overall is not). A positive NRI Cases suggest the additional variable increases true positive rates; a positive NRI controls suggest the additional variable reduces false positive rate by that amount. A negative NRI Cases suggest the additional variable reduces TPR by that amount. A negative NRI Controls suggest the additional variable increases false positive rates by that amount.

### Categorization of Protein and Genetic Scores and Association with CVD Risk

We categorized the protein and genetic scores and considered associations between the categorized scores and CVD risk to identify individuals at a high risk for CVD [Table 5]. For the genetic score model, we adjusted for the first four PCs. When the protein score was dichotomized as those individuals with scores in the top 25% versus those with scores in the bottom 75%, we found that those with scores in the top 25% were 3.9 times more likely to develop CVD compared with those with scores in the bottom 75%. Individuals with a genetic score in the top 25% were 7.3 times more likely to develop CVD compared with individuals with a score in the bottom 75%. Furthermore, individuals with a protein score and genetic score in the top 25% were 7.9 times more likely to develop CVD compared to those with both scores that were not in the top 25%.

**Table 5:**
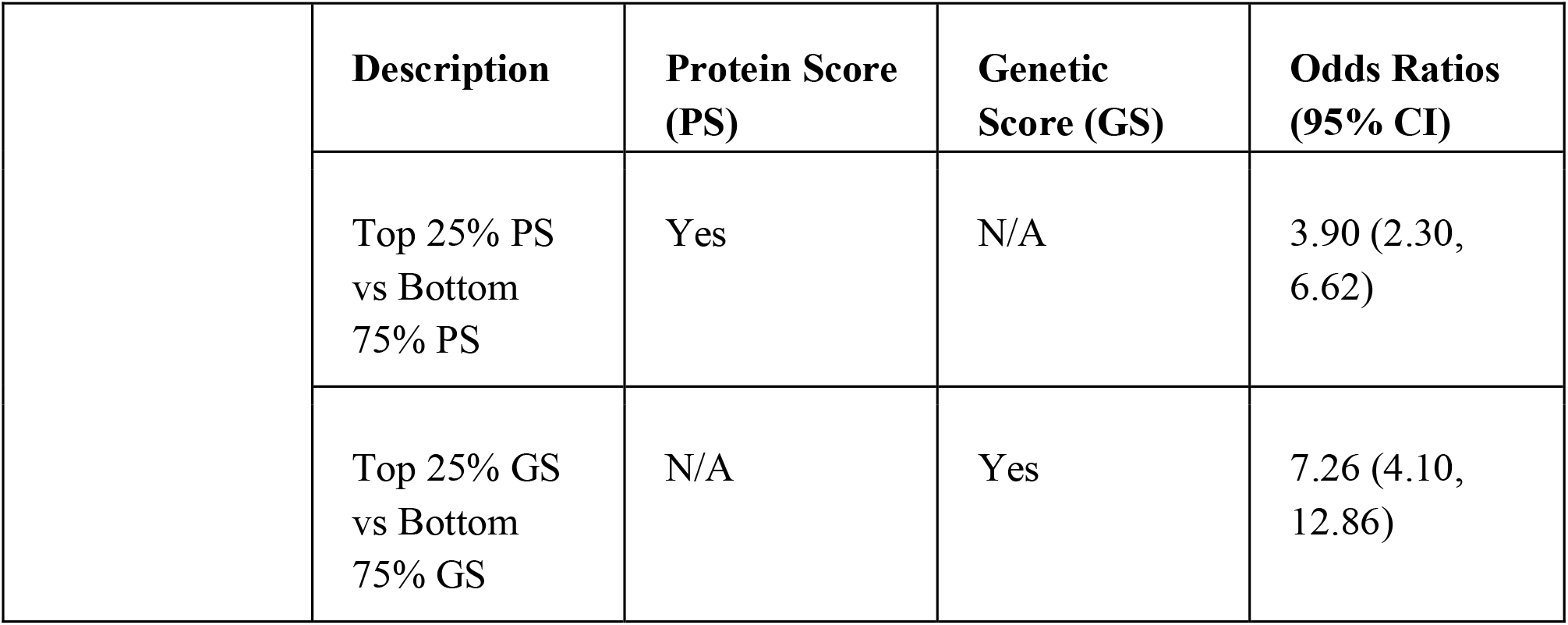
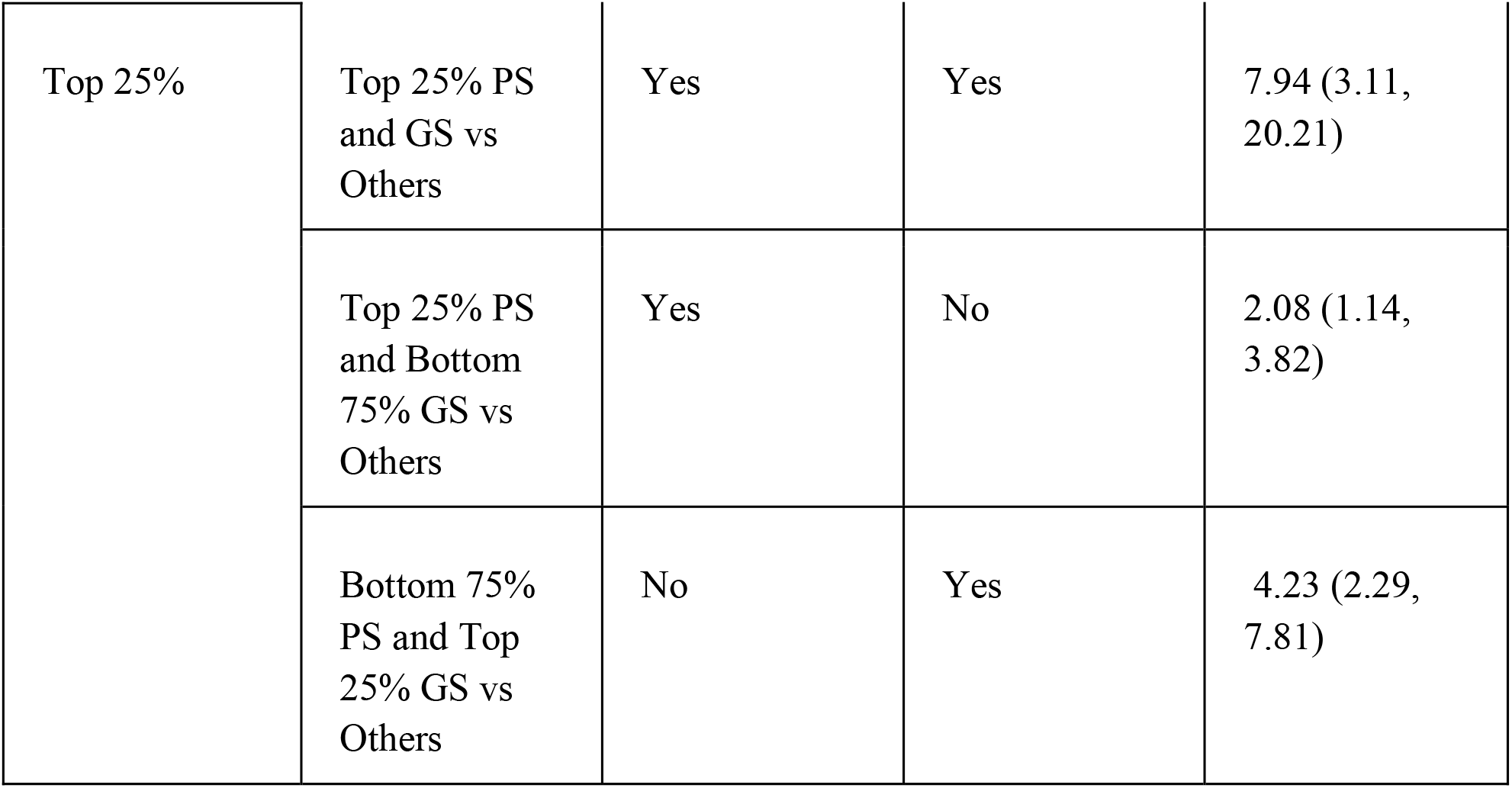
Odds ratios from categorizing protein and genetic scores. N/A means we do not include the score in the model under consideration.

|  | Description | Protein Score (PS) | Genetic Score (GS) | Odds Ratios (95% CI) |
| --- | --- | --- | --- | --- |
|  | Top 25% PS vs Bottom 75% PS | Yes | N/A | 3.90 (2.30, 6.62) |
|  | Top 25% GS vs Bottom 75% GS | N/A | Yes | 7.26 (4.10, 12.86) |
| Top 25% | Top 25% PS and GS vs Others | Yes | Yes | 7.94 (3.11, 20.21) |
|  | Top 25% PS and Bottom 75% GS vs Others | Yes | No | 2.08 (1.14, 3.82) |
|  | Bottom 75% PS and Top 25% GS vs Others | No | Yes | 4.23 (2.29, 7.81) |

## Discussion

We used resampling techniques and state-of-the-art statistical methods to analyze proteomics and genetic data from previous clinical trials to determine the impact of protein biomarkers and genetic variants on CVD risk in PLWH. Our results demonstrate that a protein risk score composed of 14 proteins and a genetic risk score composed of 15 genetic variants are more effective in predicting CVD than individual proteins or genetic variants and a baseline model consisting of established CVD and HIV risk factors. Furthermore, having a protein score in the top 25% compared to the bottom 75% resulted in a 3.9 times higher risk of CVD. Having a genetic score in the top 25% compared to the bottom 75% resulted in a 7.3 times higher risk of CVD. For individuals with both protein and gene score in the top 25% compared to others, the risk for CVD was 7.9 times higher.

Six of the 14 proteins (IL6, CCL11, SCGB3A2, GT, PLA2G7, HGF) have been linked to CVD in PLWH and were included in an 8-protein score model that improved CVD prediction^24^, also using the same data we used here. Previous studies have reported an association of IL6 with CVD in both PLWH and the general population^25–27^. HGF and PLA2G7 have both been associated with coronary heart disease in the general population^28,29,30,31^, and more recently in PLWH^32^. One novel protein is CLEC6A, which has not been extensively studied in CVD research for both PLWH and the general population. C-type lectin domain containing 6A (CLEC6A) is a protein that is encoded in humans by the CLEC6A gene. A recent Mendelian randomization analysis in PLWH that used the same cohort we used identified CLEC6A as potentially causally related to CVD^32^. IGFBP7, a potentially novel protein for PLWH, has been suggested as a marker of cellular senescence, insulin resistance, and atherosclerosis. A study found that IGFBP7 is up regulated in PLWH with fibrosis stages 2 ⁄ 3 compared to 0/1^33^. Our data suggest that IGFBP7 may be associated with CVD in PLWH. Another novel protein is GT, a member of the family of fatty acid-binding proteins, and is suggested to play an important role in metabolic function^34^. SCGB3A2 has been found in a case-control study in a Korean population to contribute to asthma susceptibility^35^ and was recently found to be potentially causally related to CVD in PLWH^32^. CCL18 and CCL11, two chemokines of the same family, have been implicated in CVD and HIV^36,37,38^. Higher levels of CCL11 have been linked to CD4+ T-cells loss^39^.

Combining CVD, HIV-related factors, genetics, and protein scores resulted in the most powerful discrimination, with an AUC of 0.86, NRI for cases of 0.53 and NRI for controls of 0.55. In a subgroup analysis restricted to individuals on ART at baseline, the genetic and protein scores were again able to differentiate between CVD cases and control. Our research builds on prior studies that created a protein score and studied the association between the score and CVD risk. In our previous work that used the same cohort^24^ but focused on only proteins, we demonstrated an 8-protein risk score combined with HIV and CVD risk factors predicted CVD risk more accurately than HIV and CVD risk factors. In the current work, we combined genetic and proteomics data, and we observed a 21.3% improvement in risk prediction compared to the 5.8% improvement in AUC in our previous work. Our AUC of 0.74 is comparable to the AUC shown in a 9-protein risk score developed and validated for coronary heart disease in the general population^40^ that went from 0.64 for the refit Framingham to 0.71 for the refit Framingham plus the 9-protein risk score, representing a 10.9% improvement in risk prediction.

Polygenic risk scores have been developed in the literature to better characterize CVD risk in the general population. However, their utility in the clinical setting is questioned, as they often require millions of SNPs to be measured, with many of these SNPs not falling in protein coding regions. This work focused on variants of genes associated with the inflammation pathway. Our proteo-genomics approach identified 15 genetic variants that, in addition to HIV and CVD factors, discriminated better between PLWH with and without CVD compared to a baseline model of CVD and HIV factors. We observed an AUC that went from 0.61 to 0.83, resulting in an improvement of 36. 1% in the model performance. This is comparable to the AUC observed from a genetic risk score for coronary artery disease in the general population^10,41^ but with only 15 genetic variants compared to the thousands to millions of genetic variants used in previous studies. This highlights the potential utility of our genetic risk score in a clinical setting. However, more studies are needed using larger sample sizes to confirm our findings.

Functional enrichment analysis of the 14 proteins used to develop the protein score revealed a strong enrichment of inflammation-related signaling pathways, such as the pathogen-induced cytokine storm-signaling pathway^42^ and the atherosclerosis-signaling pathway. Our analysis showed that the cytokine tumor necrosis factor (TNF), which is associated with many biological processes such as inflammation, immunity, apoptosis, and lipid metabolism^43^, is predicted to be activated, with 6 of 8 proteins consistent with TNF activation. Elevated TNF levels have been associated with many autoimmune diseases and are also involved in the pathophysiology of ischemia-reperfusion injury, myocarditis, and the progression of congestive heart failure^43,44^. A recent Mendelian randomization study found evidence of possible causal associations between elevated TNF levels and a higher risk of common cardiovascular diseases, such as coronary artery disease and ischemic stroke, in the general population^44^. Our data suggest that higher levels of proteins consistent with TNF activation are associated with an increased risk of CVD in PLWH. However, more studies using HIV-positive and HIV-negative controls are needed to determine the extent (if any) of this increase in PLWH compared to those without HIV.

This paper has several strengths. First, we have employed an ethnically diverse cohort, which provides insight into underrepresented and understudied groups. However, due to the size of the sample, we were unable to explore any differences in the associations between protein and genetic risk scores and CVD based on ethnicity. Second, we have used a cohort of PLWH, who are at increased risk of CVD. By combining molecular and clinical data, this paper has identified and characterized molecular variables that contribute to the risk of CVD beyond traditional risk factors for PLWH. Third, we have used genetic and protein scores with potential clinical utility. The genetic score, which consists of 15 genetic variants, allows for targeted genotyping, and focuses research efforts on clinically meaningful variants. Furthermore, the protein score, which involves only 14 proteins, is also potentially clinically applicable, as it is easily measurable and therefore more readily available for clinical use. Finally, we have used advanced statistical methods for data integration and biomarker identification. We note that when data for a subset of proteins or SNPs are available, one could still generate the risk score associated with these subsets using the weights obtained from this work.

It is essential to consider several factors when interpreting our results. First, the small sample size (n=360) limits our ability to detect modest associations and perform statistical analyses that are specific to certain populations (e.g., racial, age, and sex groups) and different genetic risk score. Second, we did not have HIV-negative controls for comparison, so it is uncertain whether the discoveries we observe are exclusive to HIV infection, and if not, to what extent our findings differ between those with and without HIV. Third, not all people were on ART (69% on ART), so it is unclear whether our findings would change if all participants were on ART at baseline. When we limited our analysis to participants on ART at baseline (n=256), the protein and genetic scores were again statistically significant and improved the prediction beyond the baseline model. Fourth, no adjustment was made for smoking status since some studies did not measure the smoking status of participants. Given that smoking is recognized as a risk factor for CVD^45^ and the smoking rate is higher in PLWH^46,47^, future research that adjusts for smoking status at baseline is necessary to determine whether our findings are still valid. Lastly, we did not have external data to validate our findings. Studies using independent data would help confirm whether our findings hold.

## Conclusion

A panel of protein biomarkers, some new (IGFBP7, HGF) and some previously known in PLWH (CLEC6A), could help identify PLWH at higher risk of developing CVD.

If confirmed, these scores could be used with CVD and HIV-related factors to identify PLWH at risk for CVD who would benefit from proactive risk reduction strategies.

## Funding and Acknowledgments

We thank participants whose data we use for this study. Sandra Safo was partially supported by grant number 1R35GM14269 from NIH. Lillian Haine was supported by the NIH under award number T32HL129956 and in part by the NIH’s National Center for Advancing Translational Sciences (NCATS), grants TL1R002493 and UL1TR002494. This study was partially funded by funding from NIH Grants UM1-AI068641, UM1-AI120197, and U01-AI136780 (for START), NIH Grants UM1-AI068641 and U01-AI46957 (for ESPRIT), NIH Grants U01-AI042170 and U01AI046362 (for FIRST) and NIH Grants UM1-AI068641, U01-AI046362, and U01-

AI042170 (for SMART). See N Engl J Med 2015; 373:795-807 for the complete list of START investigators, N Engl J Med 2009; 361:1548-59 for the complete list of ESPRIT investigators, N Engl J Med 2006; 355:2283-96 for the complete list of SMART investigators, and Lancet 2006; 368:2125-35 for the complete list of FIRST investigators. The content is solely the responsibility of the authors and does not necessarily represent the official views of the NIH’s NCATS. START was additionally supported by the National Institute of Allergy and Infectious Diseases, National Institutes of Health Clinical Center, National Cancer Institute, National Heart, Lung, and Blood Institute, Eunice Kennedy Shriver National Institute of Child Health and Human Development, National Institute of Mental Health, National Institute of Neurological Disorders and Stroke, National Institute of Arthritis and Musculoskeletal and Skin Diseases, Agence Nationale de Recherches sur le SIDA et les Hépatites Virales (France), National Health and Medical Research Council (Australia), National Research Foundation (Denmark), Bundes ministerium für Bildung und Forschung (Germany), European AIDS Treatment Network, Medical Research Council (United Kingdom), National Institute for Health Research, National Health Service (United Kingdom), and University of Minnesota. Antiretroviral drugs were donated to the central drug repository by AbbVie, Bristol-Myers Squibb, Gilead Sciences, GlaxoSmithKline/ViiV Healthcare, Janssen Scientific Affairs, and Merck.

## Data Availability

Requests for data can be made through the INSIGHT website at https://insight-trial.org. Proposals are revised by the INSIGHT Scientific Steering Committee, all individual consented to studies of genomics, which covers the IRB and consent for this project. Codes used in the analysis may be requested from the corresponding author.

## Appendix Study Group Authors

### FIRST Study Group

#### Found here

Rodger D MacArthur, Richard M Novak, Grace Peng, Li Chen, Ying Xiang, Katherine Huppler Hullsiek, Michael J Kozal, Mary van den Berg-Wolf, Christopher Henely, Barry Schmetter, Marjorie Dehlinger, for the CPCRA 058 Study Team and the Terry Beirn Community Programs for Clinical Research on AIDS (CPCRA)

#### Contributors

RDM, RMN, GP, LC, MBW, BS, and MD designed the study and wrote the study protocol. RDM, RMN, GP, LC, YX, MJK, MBW, CH, and BS implemented the study. GP, LC, YX, and KHH did the statistical analyses. RDM, RMN, MJK, and MBW were responsible for patient accrual. All authors assisted with interpretation of results and manuscript preparation.

#### Trial participants

Southern New Jersey AIDS Clinical Trials, Camden NJ, USA (K Casey, D Condoluci, D McIntyre); Wayne State University, Detroit MI, USA (L R Crane, P Chandrasekar, M Farrough); Houston AIDS Research Team, Houston TX, USA (R C Arduino, M Rodriguez-Barradas, F Visnegarwala); Richmond AIDS Consortium, Richmond, VA, USA (E Fisher, C Clark, M Pittman); North Jersey Community Research Initiative, Newark, NJ, USA (G Perez, R Roland, N Regevik); AIDS Research Alliance, Chicago, IL, USA (J P Uy, R Luskin-Hawk, M Giles); Harlem AIDS Treatment Group, New York, NY, USA (W El-Sadr, F Siegal, R Contreras); Henry Ford Hospital, Detroit, MI, USA (N P Markowitz, I Brar, L H Makohon); Denver Community Program for Clinical Research on AIDS, Denver, CO, USA (D Cohn, F Moran, R Fernandez); Louisiana Community AIDS Research Program, New Orleans, LA, USA (D Mushatt, M O Stuart, D L Dandridge); Research and Education Group, Portland OR, USA (J Sampson, D Antoniskis, J Leggett); Wide-Reaching AIDS Program, Washington DC, USA (M Turner, C Jones, D Thomas); Community Consortium of San Francisco, San Francisco CA, USA (R C Scott, P Pell); Bronx AIDS Research Consortium, Bronx, NY, USA (E E Telzak, R B Cindrich, J Shuter); Temple University, Philadelphia, PA, USA (E M Tedaldi, K Lattanzi, S Frederick); New England Program for AIDS Clinical Trials, New Haven, CT, USA (J Jensen, L Daly, L Andrews); Partners in Research New Mexico, Albuquerque, NM, USA (S B Williams, C S Nicholson, K Hammer); AIDS Research Consortium of Atlanta, Atlanta, GA, USA (B Sweeton, L Miller, M Thompson)

### ESPRIT and SILCAT Study groups

#### found HERE

**Writing Group:** Members of the writing group for the International Network for Strategic Initiatives in Global HIV Trials (INSIGHT)– Evaluation of Subcutaneous Proleukin in a Randomized International Trial (ESPRIT) Study Group and the Subcutaneous Recombinant, Human Interleukin-2 in HIV-Infected Patients with Low CD4+ Counts under Active Antiretroviral Therapy (SILCAAT) Scientific Committee (D. Abrams, M.D. [cochair], Y. Lévy, M.D. [cochair],M.H. Losso, M.D. [cochair], A. Babiker, Ph.D., G. Collins, M.S., D.A. Cooper, M.D., J. Darbyshire, M.B., Ch.B., S. Emery, Ph.D., L. Fox, M.D., F. Gordin, M.D., H.C. Lane, M.D., J.D. Lundgren, M.D., R. Mitsuyasu, M.D., J.D. Neaton, Ph.D., A. Phillips, Ph.D., J.P. Routy, M.D., G. Tambussi, M.D., and D. Wentworth, M.P.H.

**Coordinating Centers: Copenhagen:** B Aagaard, E Aragon, J Arnaiz, L Borup, B Clotet, U Dragsted, A Fau, D Gey, J Grarup, U Hengge, P Herrero, P Jansson, B Jensen, K Jensen, H Juncher, P Lopez, J Lundgren, C Matthews, D Mollerup, M Pearson, A Phillips, S Reilev, K Tillmann, S Varea. London: B Angus, A Babiker, B Cordwell, J Darbyshire, W Dodds, S Fleck, J Horton, F Hudson, Y Moraes, F Pacciarini, A Palfreeman, N Paton, N Smith, F van Hooff.

**Minneapolis:** J Bebchuk, G Collins, E Denning, A DuChene, L Fosdick, M Harrison, K Herman-Lamin, E Krum, G Larson, J Neaton, R Nelson, K Quan, S Quan, T Schultz, G Thompson, D Wentworth, N Wyman. Sydney: C Carey, F Chan, D Cooper, B Cordwell, D Courtney-Rodgers, F Drummond, S Emery, M Harrod, S Jacoby, L Kearney, M Law, E Lin, S Pett, R Robson, N Seneviratne, M Stewart, E Watts.

**Washington:** E Finley, F Gordin, A Sánchez, B Standridge, M Vjecha.

**Endpoint Review Committee:** W Belloso, R Davey, D Duprez, J Gatell, J Hoy, A Lifson, C Pederson, G Perez, R Price, R Prineas, F Rhame, J Sampson, J Worley.

**Data and Safety Monitoring Board:** J Modlin, V Beral, R Chaisson, T Fleming, C Hill, K Kim, B Murray, B Pick, M Seligmann, I Weller.

**National Institute of Allergy and Infectious Disease:** K Cahill, L Fox, M Luzar, A Martinez, L McNay, J Pierson, J Tierney, S Vogel.

**International Drug Distribution (CTS Inc**., **Durham, North Carolina):** V Costas, J Eckstrand.

**Specimen Repository (SAIC Frederick, Inc.):** S Brown.

**Clinical Sites for ESPRIT and/or SILCAAT: Argentina:** L Abusamra, E Angel, S Aquilia, W Belloso, J Benetucci, V Bittar, E Bogdanowicz, P Cahn, A Casiro, J Contarelli, J Corral, L Daciuk, D David, W Dobrzanski, A Duran, J Ebenrstejin, I Ferrari, D Fridman, V Galache, G Guaragna, S Ivalo, A Krolewiecki, I Lanusse, H Laplume, M Lasala, R Lattes, J Lazovski, G

Lopardo, M Losso, L Lourtau, S Lupo, A Maranzana, C Marson, L Massera, G Moscatello, S Olivia, I Otegui, L Palacios, A Parlante, H Salomon, M Sanchez, C Somenzini, C Suarez, M Tocci, J Toibaro, C Zala. **Australia:** S Agrawal, P Ambrose, C Anderson, J Anderson, D Baker, K Beileiter, K Blavius, M Bloch, M Boyle, D Bradford, P Britton, P Brown, T Busic, A Cain, L Carrall, S Carson, I Chenoweth, J Chuah, F Clark, J Clemons, K Clezy, D Cooper, P Cortissos, N Cunningham, M Curry, L Daly, C D’Arcy-Evans, R Del Rosario, S Dinning, P Dobson, W Donohue, N Doong, C Downs, E Edwards, S Edwards, C Egan, W Ferguson, R Finlayson, C Forsdyke, L Foy, T Franic, A Frater, M French, D Gleeson, J Gold, P Habel, K Haig, S Hardy, R Holland, J Hoy, J Hudson, R Hutchison, N Hyland, R James, C Johnston, M Kelly, M King, K Kunkel, H Lau, J Leamy, D Lester, J Leung, A Lohmeyer, K Lowe, K MacRae, C Magness, O Martinez, H Maruszak, N Medland, S Miller, J Murray, P Negus, R Newman, M Ngieng, C Nowlan, J Oddy, N Orford, D Orth, J Patching, M Plummer, S Price, R Primrose, I Prone, H Ree, C Remington, R Richardson, S Robinson, G Rogers, J Roney, N Roth, D Russell, S Ryan, J Sarangapany, T Schmidt, K Schneider, C Shields, C Silberberg, D Shaw, J Skett, D Smith, T Meng Soo, D Sowden, A Street, B Kiem Tee, Jl Thomson, S Topaz, R Vale, C Villella, A Walker, A Watson, N Wendt, L Williams, D Youds. **Austria:** A Aichelburg, P Cichon, B Gemeinhart, A Rieger, B Schmied, V Touzeau-Romer, N Vetter. **Belgium:** R Colebunders, N Clumeck, A DeRoo, K Kabeya, E O’Doherty, S de Wit. **Brazil:** C De Salles Amorim, C Basso, S Flint, E Kallas, G Levi, D Lewi, L Pereira Jr, M da Silva, T Souza, A Toscano. **Canada (CTN):** J Angel, M Arsenault, M Bast, B Beckthold, P Bouchard, I Chabot, R Clarke, J Cohen, P Coté, M Ellis, C Gagne, J Gill, M Houde, B Johnston, N Jubinville, C Kato, N Lamoureux, G Larson, J LatendrePaquette, A Lindemulder, A McNeil, N McFarland, J Montaner, C Morrisseau, R O’Neill, G Page, A Piche, B Pongracz, H Preziosi, L Puri, A Rachlis, E Ralph, I Raymond, D Rouleau, JP Routy, R Sandre,T Seddon, S Shafran, C Sikora, F Smaill, D Stromberg, S Trottier, S Walmsley, K Weiss, K Williams, D Zarowny. **Denmark:** B Baadegaard, Å Bengaard Andersen, K Boedker, P Collins, J Gerstoft, L Jensen, H Moller, P Lehm Andersen, I Loftheim, L Mathiesen, H Nielsen, N Obel, C Pedersen, D Petersen, L Pors Jensen, F Trunk Black. **France (ANRS):** JP Aboulker, A Aouba, M Bensalem, H Berthe, C Blanc, M Bloch, D Bornarel, O Bouchaud, F Boue, E Bouvet, C Brancon, S Breaud, D Brosseau, A Brunet, C Capitant, C Ceppi, C Chakvetadze, C Cheneau, JM Chennebault, P De Truchis, AM Delavalle, JF Delfraissy, P Dellamonica, AM Delavalle, C Dumont, N Edeb, G Fabre, S Ferrando, A Foltzer, V Foubert, JA Gastaut, J Gerbe, PM Girard, C Goujard, B Hoen, P Honore, H Hue,T Hynh, C Jung, S Kahi, C Katlama, JM Lang, V Le Baut, B Lefebvre, N Leturque, Y Lévy, J Loison, G Maddi, A Maignan, C Majerholc, C de Boever, JL Meynard, C Michelet, C Michon, M Mole, E Netzer, G Pialoux, I Poizot-Martin, F Raffi, M Ratajczak, I Ravaux, J Reynes, D Salmon-Ceron, M Sebire, A Simon, L Tegna, D TisneDessus, C Tramoni, JP Viard, M Vidal, C Viet-Peaucelle, L Weiss, A Zeng, D Zucman. **Germany:** A Adam, K Arastéh, G Behrens, F Bergmann, M Bickel, D Bittner, J Bogner, N Brockmeyer, N Darrelmann, M Deja, M Doerler, S Esser, G Faetkenheuer, S Fenske, S Gajetzki, D Gey, F Goebel, D Gorriahn, E Harrer, T Harrer, H Hartl, M Hartmann, S Heesch, W Jakob, H Jäger, H Klinker, G Kremer, C Ludwig, K Mantzsch, S Mauss, A Meurer, A Niedermeier, N Pittack, A Plettenberg, A Potthoff, M Probst, M Rittweger, J Rockstroh, B Ross, J Rotty, E Rund, T Ruzicka, Rt Schmidt, G Schmutz, E Schnaitmann, D Schuster, T Sehr, B Spaeth, S Staszewski, HJ Stellbrink, C Stephan, T Stockey, A Stoehr, K Tillmann, A Trein, T Vaeth, M Vogel, J Wasmuth, C Wengenroth, R Winzer, E Wolf. **Ireland:** F Mulcahy, Dl Reidy. **Israel:** Y Cohen, G Drora, I Eliezer, O Godo, E Kedem, E Magen, M Mamorsky, S Pollack, Z Sthoeger, H Vered, I Yust. **Italy:** F Aiuti, M Bechi, A Bergamasco, D Bertelli, R Bruno, L Butini, M Cagliuso, G Carosi, S Casari, V Chrysoula, G Cologni, V Conti, A Costantini, A Corpolongo, G D’Offizi, F Gaiottino, M Di Pietro, R Esposito, G Filice, M Francesco, E Gianelli, C Graziella, L Magenta, F Martellotta, R Maserati, F Mazzotta, G Murdaca, G Nardini, S Nozza, F Puppo, M Pogliaghi, D Ripamonti, C Ronchetti, S Rusconi, V Rusconi, P Sacchi, N Silvia, F Suter, G Tambussi, A Uglietti, M Vechi, B Vergani, F Vichi, P Vitiello. **Japan:** A Iwamoto, Y Kikuchi, N Miyazaki, M Mori, T Nakamura, T Odawara, S Oka, T Shirasaka, M Tabata, M Takano, C Ueta, D Watanabe, Y Yamamoto. **Morocco:** I Erradey, H Himmich, K Marhoum El Filali. **The Netherlands:** W Blok, R van Boxtel, K Brinkman H Doevelaar, A van Eeden, M Grijsen, M Groot, J Juttmann, M Kuipers, S Ligthart, P van der Meulen, J Lange, N Langebeek, S Ligthart, P Reiss, C Richter, M Schoemaker, L Schrijnders-Gudde, E Septer-Bijleveld, H Sprenger, J Vermeulen, R ten Kate, B van de Ven. **Norway:** J Bruun, D Kvale, A Maeland. **Poland:** E Bakowska, M Beniowski, A Boron-Kaczmarska, J Gasiorowski, A Horban, M Inglot, B Knysz, E Mularska, M Parczewski, M Pynka, W Rymer, A Szymczak. **Portugal:** M Aldir, F Antunes, C Baptista, J da Conceicao Vera, M Doroana, K Mansinho, C Raquel A dos Santos, E Valadas, I Vaz Pinto. **Singapore:** E Chia, E Foo, F Karim, PL Lim, A Panchalingam, N Paton, A Quek. **Spain:** R AlcázarCaballero, E Aragon, J Arnaiz, J Arribas, J Arrizabalaga, X de Barron, F Blanco, E Bouza, I Bravo, S Calvo, L Carbonero, I Carpena, M Castro, B Clotet, L Cortes, M del Toro, P Domingo, M Elias, J Espinosa, V Estrada, E Fernandez-Cruz, P Fernández, H Freud, M Fuster, A Garcia, G Garcia, R Garrido, J Gatell, P Gijón, J Gonzalez-García, I Gil, A González, J González-Lahoz, P López Grosso, M Gutierrez, E Guzmán, J Iribarren, M Jiménez, A Jou, J Juega, J Lopez, P Lopez, F Lozano, L Martín-Carbonero, R Mata, G Mateo, A Menasalvas, C Mirelles, J de Miguel Prieto, M Montes, A Moreno, J Moreno, V Moreno, R Muñoz, A Ocampo, E Ortega, L Ortiz, B Padilla, A Parras, A Paster, J Pedreira, J Peña, R Perea, B Portas, J Puig, F Pulido, M Rebollar, J de Rivera, V Roca, F Rodríguez-Arrondo, R Rubio, J Santos, J Sanz, G Sebastian, M Segovia, V Soriano, L Tamargo, S Varea, P Viciana, M von Wichmann. **Sweden:** G Bratt, A Hollander, P Olov Pehrson, I Petz, E Sandstrom, A Sönnerborg. **Switzerland:** E Bernasconi, V Gurtner. **Thailand:** U Ampunpong, C Auchieng, C Bowonwatanuwong, P Chanchai, P Chetchotisakd, T Chuenyan, C Duncombe, M Horsakulthai, P Kantipong, K Laohajinda, P Phanuphak, V Pongsurachet, S Pradapmook, K Ruxruntham, S Seekaew, A Sonjai, S Suwanagool, W Techasathit, S Ubolyam, J Wankoon. **United Kingdom:** I Alexander, D Dockrell, P Easterbrook, B Edwards, E Evans, M Fisher, R Fox, B Gazzard, G Gilleran, J Hand, L Heald, C Higgs, S Jebakumar, I Jendrulek, M Johnson, S Johnson, F Karim, G Kinghorn, K Kuldanek, C Leen, R Maw, S McKernan, L McLean, S Morris, M Murphy, S O’Farrell, E Ong, B Peters, C Stroud, M Wansbrough-Jones, J Weber, D White, I Williams, M Wiselka, T Yee. **United States:** S Adams, D Allegra, L Andrews, B Aneja, G Anstead, R Arduino, R Artz, J Bailowitz, S Banks, J Baxter, J Baum, D Benator, D Black, D Boh, T Bonam, M Brito, J Brockelman, S Brown, V Bruzzese, A Burnside Jr., V Cafaro, K Casey, L Cason, G Childress, Cl Clark, D Clifford, M Climo, D Cohn, P Couey, H Cuervo, R Davey Jr, S Deeks, M Dennis, M Diaz-Linares, D Dickerson, M Diez, J Di Puppo, P Dodson, D Dupre, R Elion, K Elliott, W El-Sadr, M Estes, J Fabre, M Farrough, J Flamm, S Follansbee, C Foster, C Frank, J Franz, G Frechette, G Freidland, J Frische, L Fuentes, C Funk, C Geisler, K Genther, M Giles, M Goetz, M Gonzalez, C Graeber, F Graziano, D Grice, B Hahn, C Hamilton, S Hassler, A Henson, S Hopper, M John, L Johnson, M Johnson, R Johnson, R Jones, J Kahn, M Kelly, N Klimas, M Kolber, S Koletar, A Labriola, R Larsen, F Lasseter, M Lederman, T Ling, T Lusch, R MacArthur, C Machado, L Makohon, J Mandelke, S Mannheimer, N Markowitz, M Martínez, N Martinez, M Mass, H Masur, D McGregor, D McIntyre, J McKee, D McMullen, M Mettinger, S Middleton, J Mieras, D Mildvan, P Miller, T Miller, V Mitchell, R Mitsuyasu, A Moanna, C Mogridge, F Moran, R Murphy, D Mushatt, R Nahass, D Nixon, S O’Brien, J Ojeda, P Okhuysen, M Olson, J Osterberger, W Owen, Sr. S Pablovich, S Patel, G Perez, G Pierone Jr., R Poblete, A Potter, E Preston, C Rappoport, N Regevik, M Reyelt, F. Rhame, L Riney, M Rodriguez-Barradas, M Rodriguez, Milagros Rodriguez, J Rodriguez, R Roland, C Rosmarin-DeStefano, W Rossen, J Rouff, M Saag, J Sampson, S Santiago, J Sarria, S Wirtz, U Schmidt, C Scott, A Sheridan, A Shin, S Shrader, G Simon, D Slowinski, K Smith, J Spotkov, C Sprague, D States, C Suh, J Sullivan, K Summers, B Sweeton, V Tan, T Tanner, E Tedaldi, Z Temesgen, D Thomas, M Thompson, C Tobin, N Toro, W Towner, K Upton, J Uy, S Valenti, C van der Horst, J Vita, J Voell, J Walker, T Walton, K Wason, V Watson, A Wellons, J Weise, M White, T Whitman, B Williams, N Williams, J Windham, M Witt, K Workowski, G Wortmann, T Wright, C Zelasky, B Zwickl.

### SMART Study Group

#### Found here

**Writing Group:** W.M. El-Sadr [cochair], Harlem Hospital Center and Columbia University, New York; J.D. Lundgren [cochair], Hvidovre University Hospital, Denmark; J.D. Neaton [cochair], University of Minnesota, Minneapolis; F. Gordin, Washington Veterans Affairs Medical Center, Washington, DC; D. Abrams, University of California, San Francisco; R.C. Arduino, University of Texas Medical School, Houston; A. Babiker, Medical Research Council, London; W. Burman, Denver Public Health Department; N. Clumeck, Centre Hospitalier Universitaire SaintPierre, Brussels; C.J. Cohen, Community Research Initiative of New England, Boston; D. Cohn, Denver Public Health Department; D. Cooper, National Centre in HIV Epidemiology and Clinical Research, Sydney; J. Darbyshire, Medical Research Council, London; S. Emery, National Centre in HIV Epidemiology and Clinical Research, Sydney; G. Fätkenheuer, University Hospital, Cologne; B. Gazzard, Medical Research Council, London; B. Grund, University of Minnesota, Minneapolis; J. Hoy, National Centre in HIV Epidemiology and Clinical Research, Melbourne; K. Klingman, National Institute of Allergy and Infectious Diseases, Bethesda, MD;

M. Losso, Hospital General de Agudos J.M. Ramos Mejia, Buenos Aires; N. Marko witz, Henry Ford Hospital, Detroit; J. Neu haus, University of Minnesota, Minneapolis; A. Phillips, Royal Free Hospital School of Medicine, London; and C. Rappoport, University of California, San Francisco

##### SMART study group are as follows

**Community Programs for Clinical Research on AIDS Chair’s Office and Operations Center** — F. Gordin (group leader), E. Finley, D. Dietz, C. Chesson, M. Vjecha, B. Standridge, B. Schmetter, L. Grue, M. Willoughby, A. Demers;

**Regional Coordinating Centers** — Copenhagen — J.D. Lundgren, A. Phillips, U.B. Dragsted, K.B. Jensen, A. Fau, L. Borup, M. Pearson, P.O. Jansson, B.G. Jensen, T.L. Benfield; London J.H. Darbyshire, A.G. Babiker, A.J. Palfreeman, S.L. Fleck, Y. Collaco-Moraes, B. Cordwell, W. Dodds, F. van Hooff, L. Wyzydrag; Sydney — D.A. Cooper, S. Emery, F.M. Drummond, S.A. Connor, C.S. Satchell, S. Gunn, S. Oka, M.A. Delfino, K. Merlin, C. McGinley;

**Statistical and Data Management Center** — Minneapolis — J.D. Neaton, G. Bartsch, A. DuChene, M. George, B. Grund, M. Harrison, C. Hogan (deceased), E. Krum, G. Larson, C. Miller, R. Nelson, J. Neuhaus, M.P. Roediger, T. Schultz, L. Thackeray;

**Electrocardiography Reading Center** — R. Prineas, C. Campbell;

**End Point Review Committee** — G. Perez (cochair), A. Lifson (cochair), D. Duprez, J. Hoy, C. Lahart, D. Perlman, R. Price, R. Prineas, F. Rhame, J. Sampson, J. Worley;

**NIAID Data and Safety Monitoring Board** — M. Rein (chair), R. Der Simonian (executive secretary), B.A. Brody, E.S. Daar, N.N. Dubler, T.R. Fleming, D.J. Freeman, J.P. Kahn, K.M.

Kim, G. Medoff, J.F. Modlin, R. Moellering, Jr., B.E. Murray, B. Pick, M.L. Robb, D.O. Scharfstein, J. Sugarman, A. Tsiatis, C. Tuazon, L. Zoloth; NIAID — K. Klingman, S. Lehrman;

**SMART Clinical Site Investigators (numbers of enrolled patients are in parentheses)— Argentina** (147) — J. Lazovski, W.H. Belloso, M.H. Losso, J.A. Benetucci, S. Aquilia, V. Bittar,

E.P. Bogdanowicz, P.E. Cahn, A.D. Casiró, I. Cassetti, J.M. Contarelli, J.A. Corral, A. Crinejo, L. Daciuk, D.O. David, G. Guaragna, M.T. Ishida, A. Krolewiecki, H.E. Laplume, M.B. Lasala, L. Lourtau, S.H. Lupo, A. Maranzana, F. Masciottra, M. Michaan, L. Ruggieri, E. Salazar, M. Sánchez, C. Somenzini; **Australia** (170) — J.F. Hoy, G.D. Rogers, A.M. Allworth, J.S.C. Anderson, J. Armishaw, K. Barnes, A. Carr, A. Chiam, J.C.P. Chuah, M.C. Curry, R.L. Dever, W.A. Donohue, N.C. Doong, D.E. Dwyer, J. Dyer, B. Eu, V.W. Ferguson, M.A.H. French, R.J. Garsia, J. Gold, J.H. Hudson, S. Jeganathan, P. Konecny, J. Leung, C.L. McCormack, M. McMurchie, N. Medland, R.J. Moore, M.B. Moussa, D. Orth, M. Piper, T. Read, J.J. Roney, N. Roth, D.R. Shaw, J. Silvers, D.J. Smith, A.C. Street, R.J. Vale, N.A. Wendt, H. Wood, D.W. Youds, J. Zillman; **Austria** (16) — A. Rieger, V. Tozeau, A. Aichelburg, N. Vetter; **Belgium** (95) — N. Clumeck, S. Dewit, A. de Roo, K. Kabeya, P. Leonard, L. Lynen, M. Moutschen, E. O’Doherty; **Brazil** (292) — L.C. Pereira, Jr., T.N.L. Souza, M. Schechter, R. Zajdenverg, M.M.T.B. Almeida, F. Araujo, F. Bahia, C. Brites, M.M. Caseiro, J. Casseb, A. Etzel, G.G. Falco, E.C.J. Filho, S.R. Flint, C.R. Gonzales, J.V.R. Madruga, L.N. Passos, T. Reuter, L.C. Sidi, A.L.C. Toscano; **Canada** (102) — D. Zarowny, E. Cherban, J. Cohen, B. Conway, C. Dufour, M. Ellis, A. Foster, D. Haase, H. Haldane, M. Houde, C. Kato, M. Klein, B. Lessard, A. Martel, C. Martel, N. McFarland, E. Paradis, A. Piche, R. Sandre, W. Schlech, S. Schmidt, F. Smaill, B. Thompson, S. Trottier, S. Vezina,S. Walmsley; **Chile** (49) — M.J. Wolff Reyes, R. Northland; Denmark (19) — L. Ostergaard, C. Pedersen, H. Nielsen, L. Hergens, I.R. Loftheim, K.B. Jensen; **Estonia** (5) — M. Raukas, K. Zilmer; **Finland** (21) — J. Justinen, M. Ristola; **France** (272) — P.M. Girard, R. Landman, S. Abel, S. Abgrall, K. Amat, L. Auperin, R. Barruet, A. Benalycherif, N. Benammar, M. Bensalem, M. Bentata, J.M. Besnier, M. Blanc, O. Bouchaud, A. Cabié, P. Chavannet, J.M. Chennebault, S. Dargere, X. de la Tribonniere, T. Debord, N. Decaux, J. Delgado, M. Dupon, J. Durant, V. Frixon-Marin, C. Genet, L. Gérard, J. Gilquin, B. Hoen, V. Jeantils, H. Kouadio, P. Leclercq, J.-D. Lelièvre, Y. Levy, C.P. Michon, P. Nau, J. Pacanowski, C. Piketty, I. Poizot-Martin, I. Raymond, D. Salmon, J.L. Schmit, M.A. Serini, A. Simon, S. Tassi, F. Touam, R. Verdon, P. Weinbreck, L. Weiss, Y. Yazdanpanah, P. Yeni; **German**y (215) — G. Fätkenheuer, S. Staszewski, F. Bergmann, S. Bitsch, J.R. Bogner, N. Brockmeyer, S. Esser, F.D. Goebel, M. Hartmann, H. Klinker, C. Lehmann, T. Lennemann, A. Plettenberg, A. Potthof, J. Rockstroh, B. Ross, A. Stoehr, J.C. Wasmuth, K. Wiedemeyer, R. Winzer; **Greece** (95) — A. Hatzakis, G. Touloumi, A. Antoniadou, G.L. Daikos, A. Dimitrakaki, P. Gargalianos-Kakolyris, M. Giannaris, A. Karafoulidou, A. Katsambas, O. Katsarou, A.N. Kontos, T. Kordossis, M.K. Lazanas, P. Panagopoulos, G. Panos, V. Paparizos, V. Papastamopoulos, G. Petrikkos, H. Sambatakou, A. Skoutelis, N. Tsogas, G. Xylomenos; **Ireland** (2) — C.J. Bergin, B. Mooka; **Israel** (13) — S. Pollack, M.G. Mamorksy, N. Agmon-Levin, R. Karplus, E. Kedem, S. Maayan, E. Shahar, Z. Sthoeger, D. Turner, I. Yust; **Italy** (88) — G. Tambussi, V. Rusconi, C. Abeli, M. Bechi, A. Biglino, S. Bonora, L. Butini, G. Carosi, S. Casari, A. Corpolongo, M. De Gioanni, G. Di Perri, M. Di Pietro, G. D’Offizi, R. Esposito, F. Mazzotta, M. Montroni, G. Nardini, S. Nozza, T. Quirino, E. Raise; **Japan** (15) — M. Honda, M. Ishisaka; **Lithuania** (4) — S. Caplinskas, V. Uzdaviniene; **Luxembourg** (3) — J.C. Schmit, T. Staub; **Morocco** (42) — H. Himmich, K. Marhoum El Filali; **New Zealand** (7) — G.D. Mills, T. Blackmore, J.A. Masters, J. Morgan, A. Pithie; **Norway** (17) — J. Brunn, V. Ormasssen; **Peru** (57) — A. La Rosa, O. Guerra, M. Espichan, L. Gutierrez, F. Mendo, R. Salazar; **Poland** (54) — B. Knytz, A. Horban, E. Bakowska, M. Beniowski, J. Gasiorowski, J. Kwiatkowski; **Portugal** (73) — F. Antunes, R.S. Castro, M. Doroana, A. Horta, K. Mansinho, A.C. Miranda, I.V. Pinto, E. Valadas, J. Vera; **Russia** (17) — A. Rakhmanova, E. Vinogradova, A. Yakovlev, N. Zakharova; **South Africa** (26) — R. Wood, C. Orrel; **Spain** (100) — J. Gatell, J.A. Arnaiz, R. Carrillo, B. Clotet, D. Dalmau, A. González, Q. Jordano, A. Jou, H. Knobel, M. Larrousse, R. Mata, J.S. Moreno, E. Oretaga, J.N. Pena, F. Pulido, R. Rubio, J. Sanz, P. Viciana; **Switzerland** (91) — B. Hirschel, R. Spycher, M. Battegay, E. Bernasconi, S. Bottone, M. Cavassini, A. Christen, C. Franc, H.J. Furrer, A. Gayet-Ageron, D. Genné, S. Hochstrasser, L. Magenta, C. Moens, N. Müller, R. Nüesch; **Thailand** (159) — P. Phanuphak, K. Ruxrungtham, W. Pumpradit, P. Chetchotisakd, S. Dangthongdee, S. Kiertiburanakul, V. Klinbuayaem, P. Mootsikapun, S. Nonenoy, B. Piyavong, W. Prasithsirikul, P. Raksakulkarn; **United Kingdom** (214) — B.G. Gazzard, J.G. Ainsworth, J. Anderson, B.J. Angus, T.J. Barber, M.G. Brook, C.D. Care, D.R. Chadwick, M. Chikohora, D.R. Churchill, D. Cornforth, D.H. Dockrell, P.J. Easterbrook, P.A. Fox, R. Fox, P.A. Gomez, M.M. Gompels, G.M. Harris, S. Herman, A.G.A. Jackson, S.P.R. Jebakumar, M.A. Johnson, G.R. Kinghorn, K.A. Kuldanek, N. Larbalestier, C. Leen, M. Lumsden, T. Maher, J. Mantell, R. Maw, S. McKernan, L. McLean, S. Morris, L. Muromba, C.M. Orkin, A.J. Palfreeman, B.S. Peters, T.E.A. Peto, S.D. Portsmouth, S. Rajamanoharan, A. Ronan, A. Schwenk, M.A. Slinn, C.J. Stroud, R.C. Thomas, M.H. Wansbrough-Jones, H.J. Whiles, D.J. White, E. Williams, I.G. Williams, M. Youle; **United States** (2989) — D.I. Abrams, E.A. Acosta, S. Adams, A. Adamski, L. Andrews, D. Antoniskis, D.R. Aragon, R. Arduino, R. Artz, J. Bailowitz, B.J. Barnett, C. Baroni, M. Barron, J.D. Baxter, D. Beers, M. Beilke, D. Bemenderfer, A. Bernard, C.L. Besch, M.T. Bessesen, J.T. Bethel, S. Blue, J.D. Blum, S. Boarden, R.K. Bolan, J.B. Borgman, I. Brar, B.K. Braxton, U.F. Bredeek, R. Brennan, D.E. Britt, J. Brockelman, S. Brown, V. Bruzzese, D. Bulgin-Coleman, D.E. Bullock, V. Cafaro, B. Campbell, S. Caras, J. Carroll, K.K. Casey, F. Chiang, G. Childress, R.B. Cindrich, C. Clark, M. Climo, C. Cohen, J. Coley, D.V. Condoluci, R. Contreras, J. Corser, J. Cozzolino, L.R. Crane, L. Daley, D. Dandridge, V. D’Antuono, J.G. Darcourt Rizo Patron, J.A. DeHovitz, E. DeJesus, J. DesJardin, M. Diaz-Linares, C. Dietrich, P. Dodson, E. Dolce, K. Elliott, D. Erickson, M. Estes, L.L. Faber, J. Falbo, M.J. Farrough, C.F. Farthing, P. Ferrell-Gonzalez, H. Flynn, C. Frank, M. Frank, K.F. Freeman, N. French, G. Friedland, N. Fujita, L. Gahagan, K. Genther, I. Gilson, M.B. Goetz, E. Goodwin, F. Graziano, C.K. Guity, P. Gulick, E.R. Gunderson, C.M. Hale, K. Hannah, H. Henderson, K. Hennessey, W.K. Henry, D.T. Higgins, S.L. Hodder, H.W. Horowitz, M. Howe-Pittman, J. Hubbard, R. Hudson, H. Hunter, C. Hutelmyer, M.T. Insignares, L. Jackson, L. Jenny, M. John, D.L. Johnson, G. Johnson, J. Johnson, L. Johnson, J. Kaatz, J. Kaczmarski, S. Kagan, C. Kantor, T. Kempner, K. Kieckhaus, N. Kimmel, B.M. Klaus, N. Klimas, J.R. Koeppe, J. Koirala, J. Kopka, J.R. Kostman, M.J. Kozal, A. Kumar, A. Labriola, H. Lampiris, C. Lamprecht, K.M. Lattanzi, J. Lee, J. Leggett, C. Long, A. Loquere, K. Loveless, C.J. Lucasti, R. Luskin-Hawk, M. MacVeigh, L.H. Makohon, S. Mannheimer, N.P. Markowitz, C. Marks, N. Martinez, C. Martorell, E. McFeaters, B. McGee, D.M. McIntyre, J. McKee, E. McManus, L.G. Melecio, D. Melton, S. Mercado, E. Merrifield, J.A. Mieras, M. Mogyoros, F.M. Moran, K. Murphy, D. Mushatt, S. Mutic, I. Nadeem, J.P. Nadler, R. Nahass, D. Nixon, S. O’Brien, A. Ognjan, M. O’Hearn, K. O’Keefe, P.C. Okhuysen, E. Oldfield, D. Olson, R. Orenstein, R. Ortiz, J. Osterberger, W. Owen, F. Parpart, V. Pastore-Lange, S. Paul, A. Pavlatos, D.D. Pearce, R. Pelz, G. Perez, S. Peterson, G. Pierone, Jr., D. Pitrak, S.L. Powers, H.C. Pujet, J.W. Raaum, J. Ravishankar, J. Reeder, N. Regevik, N.A. Reilly, C. Reyelt, J. Riddell IV, D. Rimland, M.L. Robinson, A.E. Rodriguez, M.C. Rodriguez-Barradas, V. Rodriguez Derouen, R. Roland, C. Rosmarin, W.L. Rossen, J.R. Rouff, J.H. Sampson, M. Sands, C. Savini, S. Schrader, M.M. Schulte, C. Scott, R. Scott, H. Seedhom, M. Sension, A. Sheble-Hall, A. Sheridan, J. Shuter, L.N. Slater, R. Slotten, D. Slowinski, M. Smith, S. Snap, D.M. States, M. Stewart, G. Stringer, J. Sullivan, K.K. Summers, K. Swanson, I.B. Sweeton, S. Szabo, E.M. Tedaldi, E.E. Telzak, Z. Temesgen, D. Thomas, M.A. Thompson, S. Thompson, C. Ting Hong Bong, C. Tobin, J. Uy, A. Vaccaro, L.M. Vasco, I. Vecino, G.K. Verlinghieri, F. Visnegarwala, B.H. Wade, V. Watson, S.E. Weis, J.A. Weise, S. Weissman, A.M. Wilkin, L. Williams, J.H. Witter, L. Wojtusic, T.J. Wright, V. Yeh, B. Young, C. Zeana, J. Zeh; **Uruguay** (3) — E. Savio, M. Vacarezza.

### START Study Group

#### Found here

**Writing Group:** Jens D. Lundgren, M.D. Abdel G. Babiker, Ph.D. [cochair], Fred Gordin, M.D. [cochair], Sean Emery, Ph.D., Birgit Grund, Ph.D., Shweta Sharma, M.S., Anchalee Avihingsanon, M.D., David A. Cooper, M.D., Gerd Fätkenheuer, M.D., Josep M. Llibre, M.D., Jean-Michel Molina, M.D., Paula Munderi, M.D., Mauro Schechter, M.D., Robin Wood, M.D., Karin L. Klingman, M.D., Simon Collins, H. Clifford Lane, M.D., Andrew N. Phillips, Ph.D., and James D. Neaton, Ph.D. [INSIGHT PI])

In addition to writing group, the following committee members contributed to the conduct of the START trial:

**Community Advisory Board:** C. Rappoport (INSIGHT community liaison), P.D. Aagaard, S. Collins, G.M. Corbelli,N. Geffen, C. Kittitrakul, T. Maynard, M. Meulbroek, D. Munroe, M.S. Nsubuga, D. Peavey, S. Schwarze, M. Valdez.

**Substudy Chairs:** J.V. Baker, D. Duprez (arterial elasticity); A. Carr, J. Hoy (bone mineral density); M. Dolan, A.Telenti (genomics); C. Grady (informed consent); G. Matthews, J. Rockstroh (liver fibrosis progression); W.H.Belloso, J.M. Kagan (monitoring); E. Wright, B. Brew,

R.W. Price, K. Robertson, L. Cysique (neurology); K.M.Kunisaki, J.E. Connett, D.E. Niewoehner (pulmonary). Endpoint Review Committee: A. Lifson (chair), W.H.Belloso, R.T. Davey Jr., D. Duprez, J.M. Gatell, J. Hoy, C. Pedersen, R.W. Price, R. Prineas, J. Worley.

**Central Drug Repository and Drug Distribution:** K. Brekke, S. Meger, B. Baugh, J. Eckstrand, C. Gallagher, J. Myers, J. Rooney, J. Van Wyk.

**Network Laboratory Group:** J. Baxter, C. Carey, A. DuChene, E.B. Finley, M. George, J. Grarup, M. Hoover, R. Pedersen, C. Russell, B. Standridge.

**Specimen Repositories:** E. Flowers, M. Hoover, K. Smith (Advanced BioMedical Laboratories, LLC, Cinnaminson, NJ, United States); M. McGrath, S. Silver (AIDS and Cancer Specimen Resource, University of California, San Francisco, San Francisco, CA, United States).

Wake Forest ECG Reading Center, Winston-Salem, NC, United States: E.Z. Soliman, M. Barr, C. Campbell, S. Hensley, J. Hu, L. Keasler, Y. Li, T. Taylor, Z.M. Zhang.

**Division of AIDS, National Institute of Allergy and Infectious Diseases**, Bethesda, MD, United States: B. Alston-Smith, E. DeCarlo, K. Klingman, M. Proschan.

**Data and Safety Monitoring Board:** S. Bangdiwala (chair), R. Chaisson, A.R. Fleischman, C. Hill, J. Hilton, O.H.M. Leite, V.I. Mathan, B. Pick, C. Seas, P. Suwangool, G. Thimothe, F. Venter, I. Weller, P. Yeni.

**Minnesota Coordinating Center**, University of Minnesota, Minneapolis, MN, United States: J.D. Neaton, K. Brekke, G. Collins, E.T. Denning, A. DuChene, N.W. Engen, M. George, B. Grund, M. Harrison, K.H. Hullsiek, L.H. Klemme, E. Krum, G. Larson, S. Meger, R. Nelson, J. Neuhaus Nordwall, K. Quan, S.F. Quan, T. Schultz, S. Sharma, G. Thompson.

**International Coordinating Centers:** Copenhagen HIV Programme, Rigshospitalet, University of Copenhagen, Denmark: J.D. Lundgren, B. Aagaard, A.H.D. Borges, M. Eid, J. Grarup, P. Jansson, Z. Joensen, B. Nielsen, M. Pearson, R. Pedersen, A.N. Phillips; The Kirby Institute, University of New South Wales, Sydney, Australia: S. Emery, N. Berthon-Jones, C. Carey, L. Cassar, M. Clewett, D. Courtney-Rodgers, P. Findlay, S. Hough, S. Jacoby, J. Levitt, S.L. Pett, R. Robson, V. Shahamat, A. Shambrook; Medical Research Council Clinical Trials Unit at UCL, London, United Kingdom: A.G. Babiker, B. Angus, A. Arenas-Pinto, R. Bennett, N. Braimah, E. Dennis, N. Doyle, M. Gabriel, F. Hudson, B. Jackson, A. Palfreeman, N. Paton, C. Purvis, C. Russell; Veterans Affairs Medical Center, Washington, DC, United States: F. Gordin, D. Conwell, H. Elvis, E.B. Finley, V. Kan, L. Lynch, J. Royal, A. Sánchez, B. Standridge, D. Thomas, M. Turner, M.J. Vjecha.

The following investigators participated in the START study, listed by country (country lead, numbers of participants enrolled) and clinical site:

**Argentina** (M.H. Losso, n=216): CAICI (Instituto Centralizado de Assistencia e Investigación Clínica Integral), Rosario Santa Fe: S. Lupo, L. Marconi, D. Aguila; FUNCEI, Buenos Aires: G. Lopardo, E. Bissio, D. Fridman; Fundación IDEAA, Buenos Aires: H. Mingrone, E. Loiza, V. Mingrone; Hospital General de Agudos JM Ramos Mejia, Buenos Aires: M. Losso, J.M. Bruguera, P. Burgoa; Hospital Interzonal General de Agudos Dr. Diego Paroissien, Buenos Aires: E. Warley, S. Tavella; Hospital Italiano de Buenos Aires, Buenos Aires: W. Belloso, M. Sanchez; Hospital Nacional Profesor Alejandro Posadas, Buenos Aires: H. Laplumé, L. Daciuk; Hospital Rawson, Cordoba: D. David, A. Crinejo; Argentinean SCC, Fundación IBIS, Buenos Aires: G. Rodriguez-Loria, L. Doldan, A. Moricz, I. Otegui, I. Lanusse. **Australia** (J. Hoy, n=109): Burwood Road General Practice, Burwood, VIC: N. Doong, S. Hewitt; Centre Clinic, St Kilda, VIC: B.K. Tee; East Sydney Doctors, Darlinghurst, NSW: D. Baker, E. Odgers; Holdsworth House Medical Practice, Darlinghurst, NSW: S. Agrawal, M. Bloch; Melbourne Sexual Health Centre, Carlton, VIC: T.R.H. Read, S.J. Kent; Prahran Market Clinic, Prahran, VIC: H. Lau, N. Roth; Royal Adelaide Hospital, Adelaide, SA: L. Daly, D. Shaw; Royal Perth Hospital, Perth, WA: M. French, J. Robinson; Sexual Health & HIV Service - Clinic 2, Brisbane, QLD: M. Kelly, D. Rowling; St Vincent’s Hospital, Fitzroy, VIC: D.A. Cooper, A. J. Kelleher; Taylor Square Private Clinic, Surry Hills, NSW: C. Pell, S. Dinning; The Alfred Hospital, Melbourne, VIC: J. Hoy, J. Costa; Westmead Hospital, Westmead, NSW: D.E. Dwyer, P. King. **Austria** (A. Rieger, n=7): Otto-Wagner-Spital SMZ /Baumgartner Hoehe, Vienna: N. Vetter; B. Schmied; University Vienna General Hospital, Vienna: A. Rieger, V.R. Touzeau.**Belgium** (S. de Witt, n=102): Centre Hospitalier Universitaire St. Pierre (C.H.U. St. Pierre), Brussels: S. de Witt, N. Clumeck, K. Kabeya; Institute of Tropical Medicine, Antwerp: E. Cleve, E. Florence, L. van Petersen; Universitair Ziekenhuis Gasthuisberg, Leuven: H. Ceunen, E.H. van Wijngaerden; Universitaire Ziekenhuizen Gent, Gent: T. James, L. Vandekerckhove. **Brazil** (L.C. Pereira Jr., M. Schechter, n=619): Ambulatório de Imunodeficiências (LIM-56), Sao Paulo, SP: J. Casseb, E. Constantinov, M.A. Monteiro; Center for ID at UFES, Vitoria, ES: L.N. Passos, T. Reuter; Centro de Referência e Treinamento DST/AIDS, Sao Paulo, SP: S.T. Leme, J.V.R. Madruga, R.S. Nogueira; Hospital Escola Sao Francisco de Assis, Rio de Janeiro, RJ: M. Barbosa Souza, C. Beppu Yoshida, M. Dias Costa; Instituto de Infectologia Emilio Ribas, Sao Paulo, SP: R. Castro, R.Cruz, S. Ito, T.N. Lobato Souza; Instituto FIOCRUZ, Rio de Janeiro, RJ: B. Grinsztejn, V.G. Veloso, S. Wagner Cardoso; SEI Serviços Especializados em Infectología LTDA, Salvador, Bahia: F. Bahia, C. Brites, J. Correia.**Chile** (M.J. Wolff, n=76): Fundación Arriarán, Santiago: M. Wolff, R. Northland, C. Cortés.**Czech Republic** (D. Sedlacek, n=13): Faculty Hospital Na Bulovce, Prague: D. Jilich; University Hospital Plzen, Plzen: D. Sedlacek. **Denmark** (J. Gerstoft, n=33): Hvidovre University Hospital, Hvidovre: P. Collins, L. Mathiesen; Odense University Hospital, Odense: L. Hergens, C. Pedersen; Rigshospitalet, Copenhagen: J. Gerstoft, L.P. Jensen; Århus Universitetshospital, Skejby, Århus: I.R. Lofthiem, L. Østergaard.**Estonia** (K. Zilmer, n=8): West Tallinn Central Hospital Infectious Diseases, Tallinn: K. Zilmer. **Finland** (M. Ristola, n=23): Helsinki University Central Hospital, Helsinki: M. Ristola, O. Debnam. **France** (B. Hoen, n=111): CHU Côte de Nacre – Caen, Caen: R. Verdon, S. Dargere; CHU de Besançon - Hôpital Jean-Minjoz, Besancon: B. Hoen, C. Chirouze; Groupe Hospitalier Pitié-Salpêtrière, Paris: C. Katlama, M-A. Valantin; Hôpital Antoine Béclère, Clamart: F. Boue, I. Kansau; Hôpital de Bicêtre, Le Kremlin-Bicetre: C. Goujard, C. Chakvetadze; Hôpital Européen Georges Pompidou, Paris: L. Weiss, M Karmochkine; Hôpital Foch, Suresnes: D. Zucman, C. Majerholc; Hôpital Gustava Dron, Tourcoing: O.Robineau, R. Biekre; Hôpital Henri Mondor, Creteil: Y. Levy, J.D. Lelievre; Hôpital Hôtel Dieu, Paris: J.P. Viard, J Ghosn; Hôpital Saint-Antoine, Paris: J. Pacanowski, B. Lefebvre; Hôpital Saint-Louis, Paris: J.-M. Molina, L. Niedbalski, M. Previlon; French SCC, ANRS-Inserm SC10, Paris: J.P. Aboulker, C. Capitant, B. Lebas, N. Leturque, L. Meyer, E. Netzer. **Germany** (G. Fätkenheuer, n=312): EPIMED, Berlin: K. Arastéh, T. Meier; Gemeinschaftspraxis Jessen-Jessen-Stein, Berlin: C. Zedlack, H. Jessen; ICH Study Center, Hamburg: S. Heesch, C. Hoffmann; Ifi - Studien und Projekte GmbH, Hamburg: A. Plettenberg,

A. Stoehr; Johann Wolfgang Goethe - University Hospital, Frankfurt: G. Sarrach, C. Stephan; Klinik I fu□r Innere Medizin der Universität zu Köln, Cologne: G. Fätkenheuer, E. Thomas; Klinikum der Universität Mu□nchen, Munich: J.R. Bogner, I. Ott; Klinikum Dortmund GmbH, Dortmund: M. Hower, C. Bachmann; Medizinische Hochschule Hannover, Hannover: M. Stoll, R. Bieder; Medizinische Universitätsklinik - Bonn, Bonn: J. Rockstroh, B. Becker; Universitätsklinikum Du□sseldorf, Du□sseldorf: B. Jensen, C. Feind; Universitätsklinikum Erlangen, Erlangen: E. Harrer, T. Harrer; Universitätsklinikum Essen, Essen: S. Esser, H. Wiehler; Universitätsklinikum Heidelberg, Heidelberg: M. Hartmann, R. Röger; Universitätsklinikum Regensburg, Regensburg: B. Salzberger, E. Jäger; Universitätsklinikum Wu□rzburg, Wu□rzburg: H. Klinker, G. Mark; Universitätsklinikum, Hamburg-Eppendorf: J. van Lunzen, N. Zerche; German SSC, Johann Wolfgang Goethe - University Hospital, Frankfurt: V. Mu□ller, K. Tillman. **Greece** (G. Touloumi, n=101): AHEPA University Hospital, Thessaloniki Central Macedonia: S. Metallidis, O. Tsachouridou; Attikon University General Hospital, Athens: A. Papadopoulos, K. Protopapas; Evangelismos General Hospital, Athens: A. Skoutelis, V. Papastamopoulos; Hippokration University General Hospital of Athens, Athens: H. Sambatakou, I. Mariolis; Korgialenio-Benakio Hellenic Red Cross, Athens: M. K. Lazanas, M. Chini; Syngros Hospital, Athens: S. Kourkounti, V. Paparizos; Greek SCC, National Kapodistrian University of Athens, Athens: G. Touloumi, V. Gioukari, O. Anagnostou. **India** (n=91): Institute of Infectious Diseases, Pune Maharashtra: A. Chitalikar, S. Pujari; YRGCARE Medical Centre VHS, Chennai CRS: F. Beulah, N. Kumarasamy, S. Poongulali. **Ireland** (P. Mallon, n=7): Mater Misericordiae University Hospital, Dublin: P. Mallon, P. McGettrick. **Israel** (E. Kedem, n=28): Rambam Medical Center, Haifa: E. Kedem, S. Pollack; Tel Aviv Sourasky Medical Center, Tel Aviv: D. Turner. **Italy** (G. Tambussi, n=33): Lazzaro Spallanzani IRCSS, Rome: A. Antinori, R. Libertone; Ospedale San Raffaele S.r.l., Milan: G. Tambussi, S. Nozza, M.R. Parisi. **Luxembourg** (T. Staub, n=5): Centre Hospitalier de Luxembourg, Luxembourg: T. Staub, C. Lieunard. **Malaysia** (n=18): University Malaya Medical Centre, Kuala Lumpur: R.I.S.R. Azwa. **Mali** (S. Dao, n=41): SEREFO/ CESAC Mali, Bamako, Bamako: B. Baya, M. Cissé, D. Goita. **Mexico** (n=48): INCMNSZ (Instituto Nacional de Ciencias Médicas y Nutrición), Tlalpan D.F.: J. Sierra-Madero, M.E. Zghaib. **Morocco** (K.M. El Filali, n=44): University Hospital Centre Ibn Rochd, Casablanca: K.M. El Filali, I. Erradey, H.Himmich. **Nigeria** (n=50): Institute of Human Virology-Nigeria (IHVN), Garki, Abuja FCT: E. Ekong, N. Eriobu. **Norway** (V. Ormaasen, n=15): Oslo University Hospital, Ulleval, Oslo: V. Ormaasen, L. Skeie. **Peru** (A. La Rosa, n=215): Hospital Nacional Edgardo Rebagliati Martins, Lima, Lima: M. Espichan Gambirazzio, F. Mendo Urbina; Hospital Nacional Guillermo Almenara Irigoyen, Lima, Lima: R. Salazar Castro, J. Vega Bazalar; IMPACTA Salud y Educación, Lima, Lima: M.E. Guevara, R. Infante, J. Sanchez, M. Sanchez; IMPACTA San Miguel, Lima, Lima: R. Chinchay, J.R. Lama, M. Sanchez; Via Libre, Lima, Lima: E.C. Agurto, R. Ayarza, J.A. Hidalgo. **Poland** (A.J. Horban, n=68): EMC Instytut Medyczny SA, Wroclaw: B. Knysz, A. Szymczak; Uniwersytecki Szpital Kliniczny, Bialystok: R. Flisiak, A. Grzeszczuk; Wojewodzki Szpital Zakazny, Warsaw: A.J. Horban, E. Bakowska, A. Ignatowska. **Portugal** (L. Caldeira, n=67): Hospital Curry Cabral, Lisbon: F. Maltez, S. Lino; Hospital de Egas Moniz, Lisbon: K. Mansinho, T. Bapista; Hospital de Santa Maria, Lisbon: M. Doroana, A. Sequeira, L. Caldeira; Hospital Joaquim Urbano, Oporto: J. Mendez, R.S.E. Castro. **South Africa** (R. Wood, n=518): 1 Military Hospital, Pretoria Gauteng: S.A. Pitsi; Desmond Tutu HIV Centre - Cape Town, Cape Town, Western Province: R. Kaplan, N. Killa, C. Orrell, M. Rattley; Durban International Clinical Research Site, Durban, KwaZulu Natal: U.G. Lalloo, R. Mngqibisa, S. Pillay; Durban International Clinical Research Site WWH, Durban, KwaZulu Natal: J. Govender, M. John; University of Witwatersrand, Johannesburg, Gauteng: S. Badal-Faesen, N. Mwelase, M. Rassool. **Spain** (J.R. Arribas, n=234): Complejo Hospitalario Xeral Cies, Vigo Pontevedra: A.O. Hermida, F. Warncke; Hospital Clínic de Barcelona, Barcelona: J.M. Gatell, A. Gonzalez; Hospital Clínico San Carlos, Madrid: V. Estrada, M. Rodrigo; Hospital de la Santa Creu i Sant Pau, Barcelona: P. Domingo, M. Gutierrez; Hospital del Mar, Barcelona: H.J. Knobel, A. Gonzalez; Hospital La Paz, Madrid: J.R. Arribas, M. Montes Ramirez; Hospital La Princesa, Internal Medicine and Infectious Disease Service CRS, Madrid: I. de los Santo Gil, J. Sanz Sanz; Hospital Universitari Germans Trias i Pujol, Badalona: B. Clotet, J.M. Llibre, P. Cobarsi; Hospital Universitari Mutua Terrassa, Terrassa Barcelona: D. Dalmau, C. Badia; Hospital Universitario Doce de Octubre, Madrid: R. Rubio, M.M. del Amo; Hospital Universitario Príncipe de Asturias, Alcala de Henares Madrid: J. Sanz Moreno; Hospital Universitario y Politécnico La Fe, Valencia: J. López Aldeguer, S. Cuellar; Spanish SSC, Acoiba, Madrid: P. López, B. Portas, P. Herrero. **Sweden** (M. Gisslén, n=2): Sahlgrenska University Hospital, Sweden: M. Gisslén, L. Johansson; Skåne University Hospital, Malmö: C. Håkangård, K. Törqvist. **Switzerland** (H. Furrer, n=31): Bern University Hospital, Bern: H. Furrer, A. Rauch; Unite VIH/SIDA Genèva, Genèva: A.L. Calmy, B. Hirschel (retd), T Lecompte; University Hospital Basel, Basel: M. Stoeckle; University Hospital Zurich, Zu□rich: N. Muller, M. Rizo-Oberholzer; Swiss SCC, Bern University Hospital, Bern: H. Furrer, C. Bruelisauer, A. Christen, M. Lacalamita. **Thailand** (K. Ruxrungtham, n=248): Bamrasnaradura Infections Diseases Institute, Nonthaburi: W. Prasithsirikul, S. Thongyen; Chiangrai Prachanukroh Hospital, Chiang Rai: P. Kantipong, S. Khusuwan; Chonburi Regional Hospital, Chonburi: C. Bowonwatanuwong, U. Ampunpong; Chulalongkorn University Hospital, Bangkok: K. Ruxrungtham, A. Avihingsanon, W. Thiansanguankul; Khon Kaen University, Srinagarind Hospital, Khon Kaen: P. Chetchotisakd, P. Motsikapun, S. Anunnatsari; Ramathibodi Hospital, Bangkok: S. Kiertiburanakul, N. Sanmeema; Research Institute for Health Sciences (RIHES), Chiang Mai: K. Supparatpinyo, P. Sugandhavesa; Sanpatong Hospital, Chiang Mai: V. Klinbuayaem, Y. Siriwarothai; Siriraj Hospital, Bangkok Noi: W. Ratanasuwan, T Anekthananon; Thai SCC, The HIV Netherlands Australia Thailand Research Collaboration (HIV-NAT), Bangkok: W. Harnnapachewin, T. Jupimai, P. Rerksirikul. **Uganda** (P. Mugyenyi, n=349): Joint Clinical Research Center (JCRC), Kampala: P. Mugyenyi, C. Kityo, H. Mugerwa; MRC/UVRI Research Unit on AIDS, Entebbe: P. Munderi, B. Kikaire, J. Lutaakome; MRC/UVRI Research Unit on AIDS, Masaka – satellite site: Z. Anywaine. **United Kingdom** (M.A. Johnson, n=339): Barts Health NHS Trust, London: C. Orkin, J. Hand; Belfast Health and Social Care Trust (RVH), Belfast Northern Ireland: C. Emerson, S. McKernan; Birmingham Heartlands Hospital, Birmingham West Midlands: D. White, C. Stretton; Brighton and Sussex University Hospitals NHS Trust, Brighton East Sussex: M. Fisher, A. Clarke, A. Bexley; Chelsea and Westminster Hospital, London: B. Gazzard, C. Higgs, A. Jackson; Coventry and Warwickshire NHS partnership Trust, Coventry West Midlands: S. Das, A. Sahota; Gloucestershire Royal Hospital, Gloucester: A. de Burgh-Thomas, I. Karunaratne; Guy’s and St.Thomas’ NHS Foundation Trust, London: J. Fox, J.M. Tiraboschi; Imperial College Healthcare NHS Trust, London: A. Winston, B. Mora-Peris; Leicester Royal Infirmary, Leicester Leicestershire: M.J. Wiselka, L. Mashonganyika; Lewisham and Greenwich NHS Trust, London: S. Kegg, T. Moussaoui; North Manchester General Hospital, Manchester: E. Wilkins, Y. Clowes; Queen Elizabeth Hospital Birmingham, Birmingham West Midlands: J. Ross, J. Harding; Royal Berkshire Hospital, Reading Berkshire: F. Chen, S. Lynch; Royal Bournemouth Hospital, Bournemouth Dorset: E. Herieka, J. Ablorde; Royal Free London NHS Foundation Trust, London: M.A. Johnson, M. Tyrer, M. Youle; Sheffield Teaching Hospital NHS Foundation Trust, Sheffield South Yorkshire: D. Dockrell, C. Bowman; Southmead Hospital, Bristol: M. Gompels, L. Jennings; St. George’s Healthcare NHS Trust, London: P. Hay, O. Okolo; The James Cook University Hospital, Middlesbrough Cleveland: D.R. Chadwick, P. Lambert; University College London Medical School, London: I. Williams, A. Ashraf. **United States** (K. Henry, n=507): Adult Clinical Research Center, Newark, NJ: M. Paez-Quinde, S. Swaminathan; Boston University Medical Center, Boston, MA: I. Bica, M. Sullivan; Bronx-Lebanon Hospital Center, Bronx, NY: R.B. Cindrich, L.M. Vasco; Community Research Initiative of New England, Boston, MA: J. Green, H.B. Olivet; Cooper University Hospital, Camden, NJ: J. Baxter, Y. Smith; Cornell CRS, New York, NY: V. Hughes, T. Wilkin; Denver Public Health, Denver, CO: E.M. Gardner, J. Scott; Duke University, Durham, NC: J. Granholm, N. Thielman; Florida Department of Health in Orange/Sunshine Care Center, Orlando, FL: W.M. Carter, N.D. Desai; George Washington University Medical Center, Washington, DC: D.M. Parenti, G.L. Simon; Georgetown University Medical Center, Washington, DC: P. Kumar, M. Menna; Hennepin County Medical Center, Minneapolis, MN: J. Baker, R. Givot; Henry Ford Hospital, Detroit, MI: L.H. Makohon, N.P. Markowitz; Hillsborough County Health Department, Tampa, FL: M. Chow, C. Somboonwit; Infectious Disease Associates of Northwest Florida, Pensacola, FL: A.B. Brown, B.H. Wade; Lurie Children’s Hospital, Chicago, IL: J. Jensen, A. Talsky; Maternal, Child and Adolescent Center for ID/Virology USC, Alhambra, CA: A. Kovacs, L. Spencer; Mayo Clinic, Rochester, MN: S. Rizza, Z. Temesgen; Medical College of Wisconsin, Milwaukee, WI: M. Frank, S. Parker; Montefiore Medical Center, Bronx, NY: C. Rosario, J. Shuter; Mt Sinai Hospital, Chicago, IL: K. Rohit, R. Yogev; National Military Medical Center, Bethesda, MD: I. Barahona, A. Ganesan; Naval Medical Center Portsmouth NMCP, Portsmouth, VA: S. Banks, T. Lalani; Naval Medical Center San Diego NMCSD, San Diego,CA: M.F. Bavaro, S. Echols; NICE, Southfield, MI: M. Farrough, R.D. MacArthur; NIH, Bethesda, MD: R.T.Davey Jr., R. McConnell; Ohio State University, Columbus, OH: H. Harber, S.L. Koletar; Orlando Immunology Center, Orlando, FL: E. DeJesus, A.F. Garcia; Regional Center for Infectious Disease, Greensboro, NC: K. Epperson, C.N. Van Dam; San Antonio Military Health System, JBSA Fort Sam Houston, TX: J.F. Okulicz, T.J. Sjoberg; San Juan Hospital, San Juan, PR: M. Acevedo, L. Angeli; St. Jude Children’s Research Hospital, Memphis, TN: P.M. Flynn, N. Patel; Temple University, Philadelphia, PA: C. Geisler, E. Tedaldi; Texas Children’s Hospital-Baylor College of Medicine, Houston, TX: C. McMullen-Jackson, W.T. Shearer; The Research & Education Group, Portland, OR: M.D. Murphy, S.M. Sweek; Tulane University Health Sciences Center, New Orleans, LA: D. Mushatt, C. Scott; UCLA CARE 4 Families, Los Angeles, CA: M. Carter, J. Deville; UCSD Mother-Child-Adolescent HIV Program, San Diego, CA: S.A. Spector, L. Stangl; University of Florida, Department of Pediatrics, Jacksonville, FL: M.H. Rathore, K. Thoma; University of Florida, Jacksonville, FL: M. Sands, N. Wilson; University of Illinois at Chicago, Chicago, IL:. M. Novak, T. Pearson; University of Miami, Miami, FL: M.A. Kolber, T. Tanner; University of North Carolina, Chapel Hill, NC: M. Chicurel-Bayard, E. Hoffman; University of North Texas Health Science Center, Fort Worth, TX: I. Vecino, S.E. Weis; University of Puerto Rico, San Juan, PR: I. Boneta, J. Santana; University of Texas Southwestern Medical Center, Dallas, TX: M.K. Jain, M. Santos; Veterans Affairs Greater LA Healthcare System, Los Angeles, CA: M.B. Goetz, W.L. Rossen; Virginia Commonwealth University, Richmond, VA: D. Nixon, V. Watson; Wake County Human Services, Raleigh, NC: D. Currin, C. Kronk; Wake Forest University Health Sciences, Winston-Salem, NC: L. Mosley, A. Wilkin; Washington DC Veterans Administration, Washington, DC: A.M. Labriola, D.W. Thomas; Yale University School of Medicine, New Haven, CT: D. Chodkowski, G. Friedland.

