## Supplementary Material for "Proteomic and Genetic predictors and risk scores of cardiovascular diseases in persons living with HIV"

A. More on Methods

*Proteomics Data from Olink, Normalization and Quality Control*

The data are median intensity normalized, meaning that the data are adjusted to make the median value for each assay on each plate equal to the median for that of the other plates. The data are presented as normalized protein expression values (NPX) and are reported on the log_2_ scale. Four internal controls for each sample are added to monitor the quality of assay performance and the quality of each individual sample. There are two steps of quality control: 1) each sample plate with above 0.2 NPX when evaluated on the standard deviation of the internal controls pass this first step and 2) samples that deviate less than 0.3 NPX from the median value of the controls for each individual sample pass this step.

*Integrative Analysis of Genetics and Proteomics Data*

We considered a multivariate approach to investigate the associations between the genetics and proteomics data, and to determine genetic variants and proteins that are correlated and discriminate between CVD cases and controls. Proteins that appeared on multiple panels were all considered in the integrative analysis. We used the sparse integrative analysis (SIDA) approach^11^ for this purpose. SIDA is a joint association and classification method that allows to simultaneously model correlations between two or more data types (in our case the genetics and proteomics data) while allowing for discrimination between two or more groups (in our case CVD cases and controls). SIDA allows the detection of key variables that both maximize associations between the genetics and proteomics data and discriminate between CVD cases and controls. For statistical rigor, we coupled SIDA with resampling techniques to determine key molecules. We obtained 100 bootstrap training and testing datasets. On each bootstrap data, we fitted a logistic regression model of our outcome with each protein and genetic variant. The genetic variant was coded as the number of minor alleles. Molecules which were statistically significant (p-value < 0.05) were used in the SIDA algorithm to integrate the genetic and proteomics data. By embedding the univariate step into the integration step, we ensure that variables that went into SIDA discriminated between CVD cases and controls. Proteins and genes passing the univariate filtering step and were frequently selected by SIDA to discriminate between cases and controls were chosen as candidate variables for downstream analyses.

*More on Pathway Analysis of Molecules Identified in Proteo-genomic Integrative Analysis*

Regarding upstream regulators, tumor necrosis factor (TNF), lipopolysaccharide (LPS) and tetradecanoylphorbol acetate were each predicted to be activated. In particular, the cytokine TNF was predicted to be activated with a z-score of 2.077 and overlap p-value 5.65E-06. Six (HGF, LTBR, IL6, UPAR, CCL11, GT) out of 8 genes known in the literature to be upregulated by TNF were also upregulated in our dataset, which is consistent with activation of TNF. LPS was predicted to be activated with a z-score of 2.385 and overlap p-value 2.68E-05 with 6 (HGF, IL6, UPAR, CCL11, CCL18, PLA2G7) out of 8 genes consistent with activation of LPS. Tetradecanoylphorbol acetate was predicted to be activated (z-score 2.156, overlap p-value 3.27E-05) with 5 (HGF, IL6, UPAR, CCL11, CCL18) out of 6 genes consistent with activation of tetradecanoylphorbol acetate.

B. **Description of proteins used in protein score**

###### **Inflammation Panel:**

**Hepatocyte Growth Factor (HGF):**

The protein HGF and its receptor c-MET are involved in tissue repair and respond to tissue injury. HGF has been proposed as a potential clinical biomarker for CVD^45^ HGF has already been shown to be associated with stroke, CHD, atherosclerosis, and the progression of atherosclerosis in an ethnically diverse, general population^29,30^

**Fibroblast growth factor 19 (FGF19):**

FGF19, a member of the fibroblast growth factor (FGF), is a protein that in humans is encoded by the gene FG19. Proteins in the FGF family have been implicated in a variety of biological processes that includes cell growth, tissue repair, tumor growth and invasion^48^, glucose and lipid metabolism^49^. They have been suggested as therapeutic biomarkers for chronic diseases such as obesity, type 2 diabetes, cancer, and kidney and cardiovascular disease in the general population^49^.

***Immune Response Panel:***

**C-C motif chemokine 11 (CCL11):**

CCL-11 belongs to the family of cytokines implicated in immunoregulatory and inflammatory processes. Increased levels of CCL11 have been associated with coronary artery disease^38^. Higher levels of CCL11 have been linked to CD4+ T-cells loss^39^.

CLEC6A

C-type lectin domain containing 6A (CLEC6A) is a protein that is encoded in humans by the CLEC6A gene. A recent Mendelian randomization analysis in PLWH that used the same cohort we used identified CLEC6A as potentially causally related to CVD^32^.

**Interleukin-6 (IL6):**

IL6 has been extensively studied in both healthy and HIV positive populations. IL6 is a marker of inflammation and coagulation. Increased levels of plasma IL6 has been shown to associated with increased risk of CVD, atherosclerosis, and mortality in an HIV positive population even when treated with ART^25–27^.

###### **Cardiovascular 2 Panel:**

**Gastrotropin (GT):**

Gastrotropin, also known as the ileal fatty acid binding protein, (FABP6) is a member of the fatty acid-binding protein (FABPs) family, which regulates general metabolic function via FABPs central role in fatty acid transport, metabolism, and storage. FABPs have been associated with a number of diseases including cardiovascular disease and are thought to serve an integral role in metabolic function^34^. FABP6 is more specifically known to be involved in bile acid metabolism. There has been shown to be a protective association between FABP6 Thr79Met polymorphism and incident type 2 diabetes^50^.

###### **A disintegrin and metalloproteinase with thrombospondin motifs 13 (ADAMTS13**)

A disintegrin and metalloproteinase with thrombospondin motifs 13 is a protein that in human is encoded by the gene ADAMTS13. Low levels of the protein ADAMTS13 is suggested to be associated with in an increased risk of ischemic stroke, myocardial infarction and cerebrovascular disease in the general population^51^, and is thought to contribute to increased cardiovascular risk in persons with HIV^52^. In an HIV study, ADAMTS13 antigen and

**Interleukin-1 receptor-like 2 (IL1RL2**)

IL1RL2 is a protein that in humans is encoded by the **IL1RL2** gene. It is a member of the interlukin 1 receptor family, which is implicated in inflammatory diseases^53^. IL1 is suggested to contribute to the initiation, formation, growth and rupture of atherosclerosis plaques^54^.

###### **Cardiometabolic Panel**

**Platelet-activating factor acetylhydrolase (PLA2G7):**

PLA2G7 is found in both high-density lipoprotein (HDL) and low-density lipoprotein (LDL). In population studies it has been shown that overexpression of PLA2G7 is associated with increased coronary heart disease (CHD)^28,31^. It is thought that with individuals with low LDL cholesterol levels it can help predict CHD risk^28,31^.

**C-C motif chemokine 18 (CCL18):**

CCL-18 belongs to the family of cytokines implicated in immunoregulatory and inflammatory processes. CCL18 upregulation has been reported in a number of diseases, including HIV infection, atherosclerosis and pulmonary fibrosis^36,37^.

###### **Cardiovascular 3 Panel**

**Lymphotoxin beta receptor** (**LTBR**)

LTBR, also known as the tumor tumor necrosis factor receptor superfamily member 3 (TNFRSF3) is a member of the tumor necrosis factor receptor family and is implicated in apoptosis and cytokine release. This protein was found to be associated with the presence of plaque in PLWH^55^.

**urokinase-type plasminogen activator receptor or urokinase receptor (uPAR)**

uPAR is a protein encoded in humans by the plasminogen activator, urokinase receptor gene (PLAUR). It is a member of the plasminogen activator system which has been implicated in biological processes such as hemostatis and inflammation^56,57^. UPAR is predicted to be upregulated during conditions of injury and inflammation^58^. Research suggests that in persons with HIV, uPAR expression is upregulated with activation of monocytes and T lymphocytes^59^.

**Secretoglobin Family 3A Member (SCGB3A2):**

A case-control study in a Korean population on SNPs in the SCGB3A2 gene potentially contributes to susceptibility to asthma^35^. The SCGB3A2 gene is also known as the uterus globulin associated protein 1 (UGRP1) found that UGRP1 may be able to predict graves’ disease patients who develop hypothyroidism^60^.

**insulin like growth factor binding protein (IGFBP7)**

IGFBP7 (insulin like growth factor binding protein 7) is a protein which in humans is encoded by the gene IGFBP7. Studies have reported the association of IGFBP7 in a variety of cancers. Recent work suggest IGFBP7 as a marker of cellular senescence, insulin resistance and atherosclerosis^61–63^.

**Table S1: Log-odds ratios of proteins used in Ingenuity Pathway Analysis**

| **Protein** | **Log-odds ratios** | **Odds Ratios** | **P-value** |
| --- | --- | --- | --- |
| IL6 | 0.420019 | 1.521991 | 0.000195 |
| CCL11 | 0.342293 | 1.408173 | 0.002502 |
| CLEC6A | 0.342984 | 1.409146 | 0.003178 |
| HGF | 0.575619 | 1.778232 | 7.63E-06 |
| FGF19 | 0.269885 | 1.309813 | 0.015346 |
| ADAMTS13 | -0.36212 | 0.696197 | 0.063394 |
| IL1RL2 | -0.24638 | 0.781623 | 0.045265 |
| GAST | 0.224594 | 1.251814 | 0.047525 |
| CCL18 | 0.33387 | 1.396362 | 0.003123 |
| PLA2G7 | 0.274521 | 1.315901 | 0.013146 |
| LTBR | 0.453132 | 1.573232 | 0.000197 |
| PLAUR | 0.360565 | 1.43414 | 0.001672 |
| SCGB3A2 | 0.338422 | 1.402732 | 0.002127 |
| IGFBP7 | 0.488998 | 1.630682 | 6.49E-05 |

**Table S2. Coefficients from Baseline + Protein Score Model**

| **Variable** | **OR** | **LCI** | **UCI** | **p-value** |
| --- | --- | --- | --- | --- |
| Gender | 0.565 | 0.223 | 1.319 | 0.204 |
| Age | 0.986 | 0.957 | 1.014 | 0.324 |
| CVD | 5.647 | 1.574 | 27.06 | 0.014 |
| CD4 | 1 | 0.999 | 1.001 | 0.773 |
| Lipid-lowering medication | 1.217 | 0.629 | 2.318 | 0.553 |
| BP lowering medication | 1.711 | 0.835 | 3.483 | 0.139 |
| Diabetes | 0.733 | 0.251 | 2.021 | 0.555 |
| Black Race | 1.277 | 0.601 | 2.662 | 0.517 |
| Protein Score | 2.356 | 1.776 | 3.19 | < 0.001 |

**Table S3: Logistic Regression Model of CVD on Each of the Standardized Proteins Used to Develop the Protein Score (n=360)**

|  | **OR** | **SE** | **p-value** | **LCI** | **UCI** |
| --- | --- | --- | --- | --- | --- |
| Baseline + CCL11 | 1.36 | 0.127 | 0.015 | 1.064 | 1.751 |
| Baseline + CLEC6A | 1.419 | 0.13 | 0.007 | 1.106 | 1.84 |
| Baseline + IL6 | 1.439 | 0.125 | 0.004 | 1.129 | 1.847 |
| Baseline + HGF | 1.623 | 0.137 | < 0.001 | 1.252 | 2.141 |
| Baseline + FGF19 | 1.378 | 0.122 | 0.009 | 1.088 | 1.757 |
| Baseline + ADAMTS13 | 0.63 | 0.221 | 0.037 | 0.403 | 0.944 |
| Baseline + IL1RL2 | 0.761 | 0.141 | 0.053 | 0.572 | 0.993 |
| Baseline + GT | 0.747 | 0.132 | 0.027 | 0.572 | 0.959 |
| Baseline + CCL18 | 1.455 | 0.123 | 0.002 | 1.15 | 1.866 |
| Baseline + PLA2G7 | 1.356 | 0.127 | 0.017 | 1.058 | 1.746 |
| Baseline + LTBR | 1.514 | 0.129 | 0.001 | 1.185 | 1.971 |
| Baseline + UPAR | 1.373 | 0.123 | 0.01 | 1.085 | 1.759 |
| Baseline + SCGB3A2 | 1.422 | 0.12 | 0.003 | 1.127 | 1.803 |
| Baseline + IGFBP7 | 1.509 | 0.137 | 0.003 | 1.163 | 1.992 |

**Table S4: Incremental Contribution of Individual Proteins and SNPs and Protein Score to CVD Risk When Added to Baseline Model (n=360)**

| Model | AUC | LCB | UCB | NRI Cases | NRI Controls | Overall NRI |
| --- | --- | --- | --- | --- | --- | --- |
| Baseline Model | 0.612 | 0.5495 | 0.6746 |  |  |  |
| ***Baseline + Individual Proteins***  Baseline + CCL11 | 0.647 | 0.585 | 0.708 | 0.0847 | 0.048 | 0.1328 |
| Baseline + CLEC6A | 0.653 | 0.5916 | 0.7135 | 0.0847 | 0.1703 | 0.2551 |
| Baseline + IL6 | 0.679 | 0.6202 | 0.7377 | 0.0339 | 0.1965 | 0.2304 |
| Baseline + HGF | 0.671 | 0.6121 | 0.7306 | 0.136 | 0.162 | 0.297 |
| Baseline + FGF19 | 0.653 | 0.5911 | 0.7147 | 0.0339 | 0.1441 | 0.178 |
| Baseline + ADAMTS13 | 0.637 | 0.5763 | 0.6978 | 0.0847 | 0.1092 | 0.1939 |
| Baseline + IL1RL2 | 0.645 | 0.5842 | 0.7059 | 0.0678 | 0.214 | 0.2818 |
| Baseline + GT | 0.636 | 0.5745 | 0.697 | 0.271 | 0.048 | 0.319 |
| Baseline + CCL18 | 0.653 | 0.5924 | 0.7139 | 0 | 0.188 | 0.188 |
| Baseline + PLA2G7 | 0.649 | 0.5871 | 0.7108 | 0.0678 | 0.2227 | 0.2905 |
| Baseline + LTBR | 0.663 | 0.6009 | 0.7258 | 0.0508 | 0.1965 | 0.2474 |
| Baseline + UPAR | 0.661 | 0.6009 | 0.7209 | 0.153 | 0.258 | 0.41 |
| Baseline + SCGB3A2 | 0.656 | 0.5948 | 0.717 | 0.0339 | 0.2664 | 0.3003 |
| Baseline + IGFBP7 | 0.646 | 0.5826 | 0.7086 | -0.0508 | 0.1703 | 0.1195 |
| **Baseline + Protein Score** | 0.742 | 0.6865 | 0.7968 | 0.305 | 0.345 | 0.65 |
| ***Baseline + Individual SNPs***  Baseline + 4 PCs + rs11895665 | 0.649 | 0.588 | 0.711 | -0.017 | 0.223 | 0.206 |
| Baseline + 4 PCs + rs2240688 | 0.684 | 0.624 | 0.745 | 0.254 | 0.162 | 0.416 |
| Baseline + 4 PCs + rs4696483 | 0.667 | 0.607 | 0.728 | -0.169 | 0.467 | 0.298 |
| Baseline + 4 PCs + rs34308112 | 0.667 | 0.607 | 0.732 | -0.203 | 0.616 | 0.412 |
| Baseline + 4 PCs + rs10456432 | 0.694 | 0.636 | 0.751 | 0.492 | -0.013 | 0.478 |
| Baseline + 4 PCs + rs3808528 | 0.650 | 0.590 | 0.710 | 0.220 | 0.135 | 0.356 |
| Baseline + 4 PCs + rs17252559 | 0.660 | 0.600 | 0.721 | -0.169 | 0.450 | 0.280 |
| Baseline + 4 PCs + rs3816208 | 0.653 | 0.592 | 0.715 | -0.186 | 0.406 | 0.220 |
| Baseline + 4 PCs + rs9410490 | 0.660 | 0.599 | 0.721 | 0.271 | 0.118 | 0.389 |
| Baseline + 4 PCs + rs16940029 | 0.635 | 0.573 | 0.697 | 0.305 | -0.066 | 0.240 |
| Baseline + 4 PCs + rs115708227 | 0.670 | 0.612 | 0.729 | 0.746 | -0.345 | 0.401 |
| Baseline + 4 PCs + rs17053844 | 0.636 | 0.574 | 0.698 | -0.051 | 0.266 | 0.216 |
| Baseline + 4 PCs + rs80067004 | 0.646 | 0.584 | 0.708 | -0.085 | 0.284 | 0.199 |
| Baseline + 4 PCs + rs73320494 | 0.663 | 0.604 | 0.722 | 0.525 | -0.109 | 0.416 |
| Baseline + 4 PCs + rs12602462 | 0.654 | 0.592 | 0.716 | 0.254 | 0.013 | 0.267 |
| **Baseline + SNP Score + 4 PCs** | 0.829 | 0.784 | 0.874 | 0.492 | 0.467 | 0.959 |
| **Baseline + Protein Score + SNP Score + 4 PCs** | 0.858 | 0.819 | 0.897 | 0.525 | 0.546 | 1.071 |

**Table S5. Coefficients from Baseline + Protein Score Model (ART Subset)**

| **Variable** | **OR** | **LCI** | **UCI** | **p-value** |
| --- | --- | --- | --- | --- |
| Gender | 0.69 | 0.20 | 2.02 | 0.51 |
| Age | 0.99 | 0.95 | 1.02 | 0.43 |
| CVD at Baseline | 2.67 | 0.61 | 14.28 | 0.21 |
| CD4 | 1.00 | 1.00 | 1.00 | 0.83 |
| Lipid-lowering medication | 1.30 | 0.60 | 2.74 | 0.50 |
| BP lowering medication | 2.14 | 0.84 | 5.44 | 0.11 |
| Diabetes | 0.64 | 0.15 | 2.45 | 0.53 |
| Black Race | 0.99 | 0.36 | 2.53 | 0.98 |
| Protein Score | 2.27 | 1.65 | 3.20 | < 0.001 |

**Table S6. Selected Coefficients from Baseline + SNP Score + 4 PCs Model**

| **Variable** | **OR** | **LCI** | **UCI** | **p-value** |
| --- | --- | --- | --- | --- |
| Gender | 0.698 | 0.262 | 1.722 | 0.45 |
| Age | 0.99 | 0.959 | 1.022 | 0.551 |
| CVD | 9.11 | 2.101 | 50.901 | 0.006 |
| CD4 | 0.999 | 0.998 | 1.001 | 0.41 |
| Lipid-lowering medication | 1.349 | 0.646 | 2.782 | 0.42 |
| BP lowering medication | 1.202 | 0.544 | 2.637 | 0.646 |
| Diabetes | 1.082 | 0.351 | 3.192 | 0.888 |
| Black Race | 0.624 | 0.039 | 22.791 | 0.766 |
| SNP Score | 4.586 | 3.214 | 6.803 | < 0.001 |

**Table S7: Logistic Regression Model of CVD on Each of the Standardized SNPs Used to Develop the Genetic Score (n=360)**

|  | **OR** | **SE** | **p-value** | **LCI** | **UCI** |
| --- | --- | --- | --- | --- | --- |
| Baseline + 4 PCs + X2.238552973_T | 1.335 | 0.127 | 0.023 | 1.04 | 1.717 |
| Baseline + 4 PCs + X4.15970349_G | 0.671 | 0.136 | 0.003 | 0.51 | 0.871 |
| Baseline + 4 PCs + X4.154619255_T | 1.481 | 0.133 | 0.003 | 1.142 | 1.93 |
| Baseline + 4 PCs + X5.53444623_A | 1.654 | 0.126 | < 0.001 | 1.298 | 2.133 |
| Baseline + 4 PCs + X6.35671651_C | 0.552 | 0.147 | < 0.001 | 0.408 | 0.729 |
| Baseline + 4 PCs + X8.23158775_A | 1.305 | 0.122 | 0.029 | 1.028 | 1.661 |
| Baseline + 4 PCs + X8.53603859_G | 1.367 | 0.121 | 0.01 | 1.078 | 1.736 |
| Baseline + 4 PCs + X8.97614625_A | 1.391 | 0.119 | 0.006 | 1.102 | 1.762 |
| Baseline + 4 PCs + X9.92089100_C | 0.703 | 0.132 | 0.007 | 0.539 | 0.905 |
| Baseline + 4 PCs + X12.109617728_G | 1.318 | 0.145 | 0.057 | 0.993 | 1.758 |
| Baseline + 4 PCs + X12.109644408_A | 0.623 | 0.158 | 0.003 | 0.443 | 0.829 |
| Baseline + 4 PCs + X13.53625421_T | 1.248 | 0.118 | 0.061 | 0.989 | 1.579 |
| Baseline + 4 PCs + X15.101934803_G | 1.342 | 0.123 | 0.017 | 1.053 | 1.712 |
| Baseline + 4 PCs + X17.45373571_C | 0.632 | 0.148 | 0.002 | 0.467 | 0.836 |
| Baseline + 4 PCs + X17.78146016_T | 1.29 | 0.123 | 0.038 | 1.014 | 1.645 |

**Table S8. Selected Coefficients from Baseline + SNP Score + 4 PCs Model (ART Subset)**

| **Variable** | **OR** | **LCI** | **UCI** | **p-value** |
| --- | --- | --- | --- | --- |
| Gender | 0.98 | 0.28 | 3.01 | 0.97 |
| Age | 0.99 | 0.96 | 1.03 | 0.71 |
| CVD at Baseline | 5.42 | 1.03 | 33.49 | 0.05 |
| CD4 | 1.00 | 1.00 | 1.00 | 0.29 |
| Lipid-lowering medication | 1.45 | 0.62 | 3.34 | 0.38 |
| BP lowering medication | 1.48 | 0.52 | 4.22 | 0.46 |
| Diabetes | 1.08 | 0.24 | 4.50 | 0.92 |
| Black Race | 0.99 | 0.03 | 122.53 | > 0.99 |
| SNP Score | 3.82 | 2.63 | 5.81 | < 0.001 |

**Table S9. Selected Coefficients from Baseline + Protein Score + SNP Score + 4 PCs Model (ART Subset)**

| **Variable** | **OR** | **LCI** | **UCI** | **p-value** |
| --- | --- | --- | --- | --- |
| Gender | 1.22 | 0.32 | 4.12 | 0.75 |
| Age | 0.99 | 0.95 | 1.02 | 0.45 |
| CVD at Baseline | 4.28 | 0.80 | 26.59 | 0.10 |
| CD4 | 1.00 | 1.00 | 1.00 | 0.62 |
| Lipid-lowering medication | 1.53 | 0.62 | 3.77 | 0.35 |
| BP lowering medication | 2.00 | 0.69 | 6.10 | 0.22 |
| Diabetes | 0.70 | 0.13 | 3.38 | 0.67 |
| Black Race | 0.25 | 0.0057 | 48.30 | 0.52 |
| Protein Score | 2.24 | 1.55 | 3.35 | < 0.001 |
| SNP Score | 3.90 | 2.61 | 6.10 | < 0.001 |

Figures

**Figure S1 SNPs QC for Inflammation pathway**

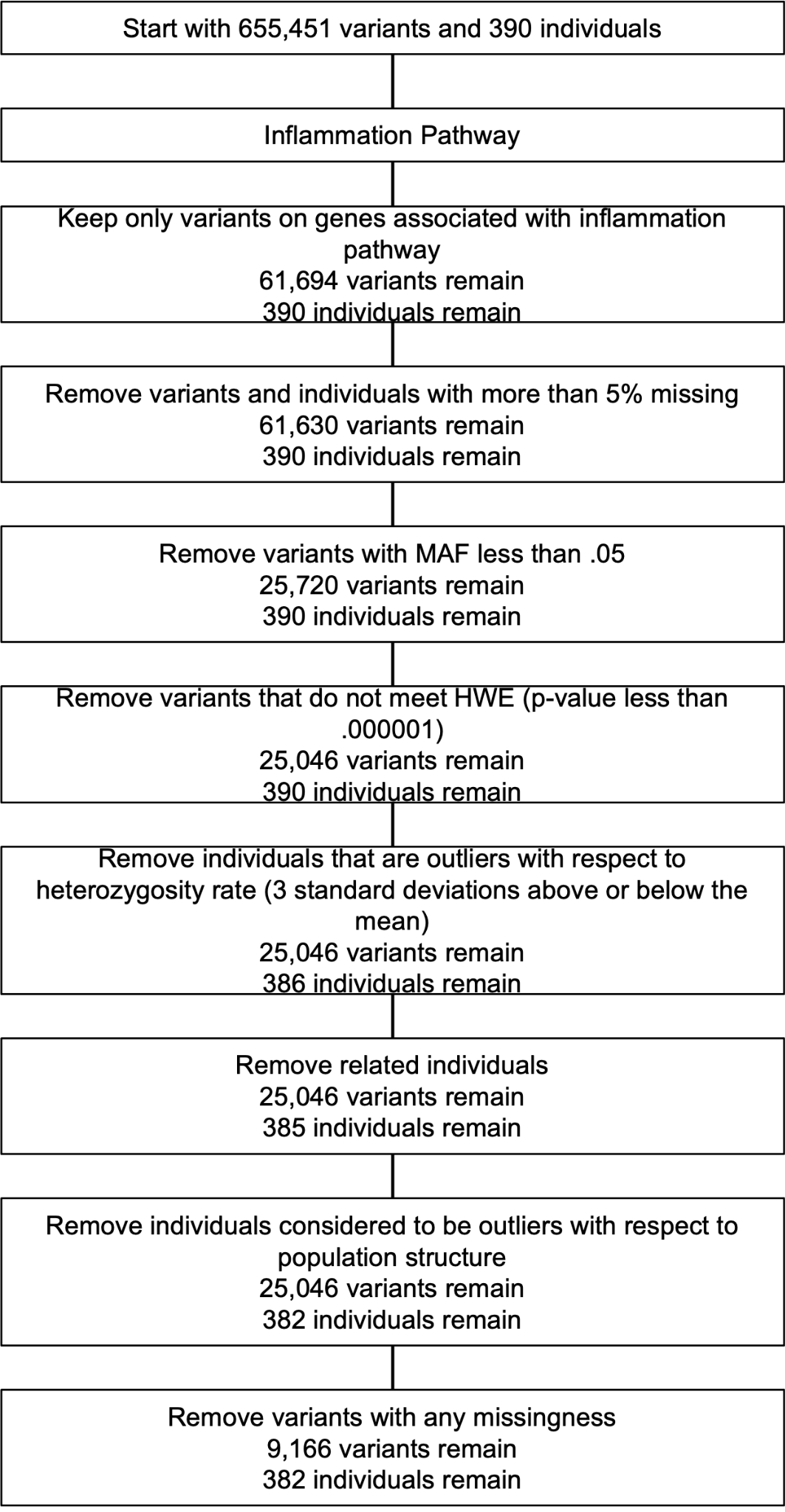

Figure S2: Correlations between SNPs and genetic variants.

**Figure S3- Unadjusted and Ancestry-adjusted polygenic risk score**

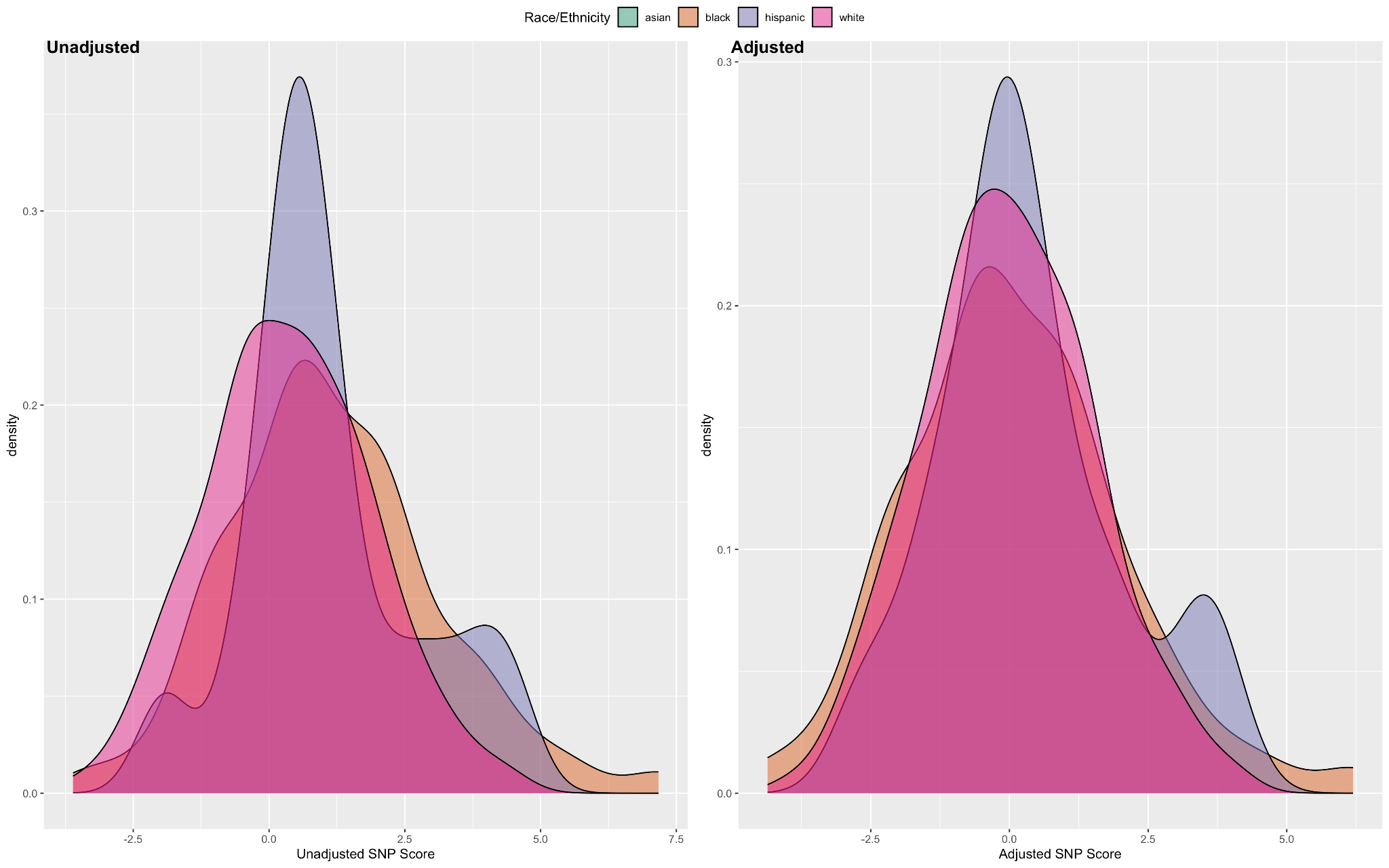

**Figure S4 Distribution of ancestry-adjusted genetic and proteomics score across demographic variables.**

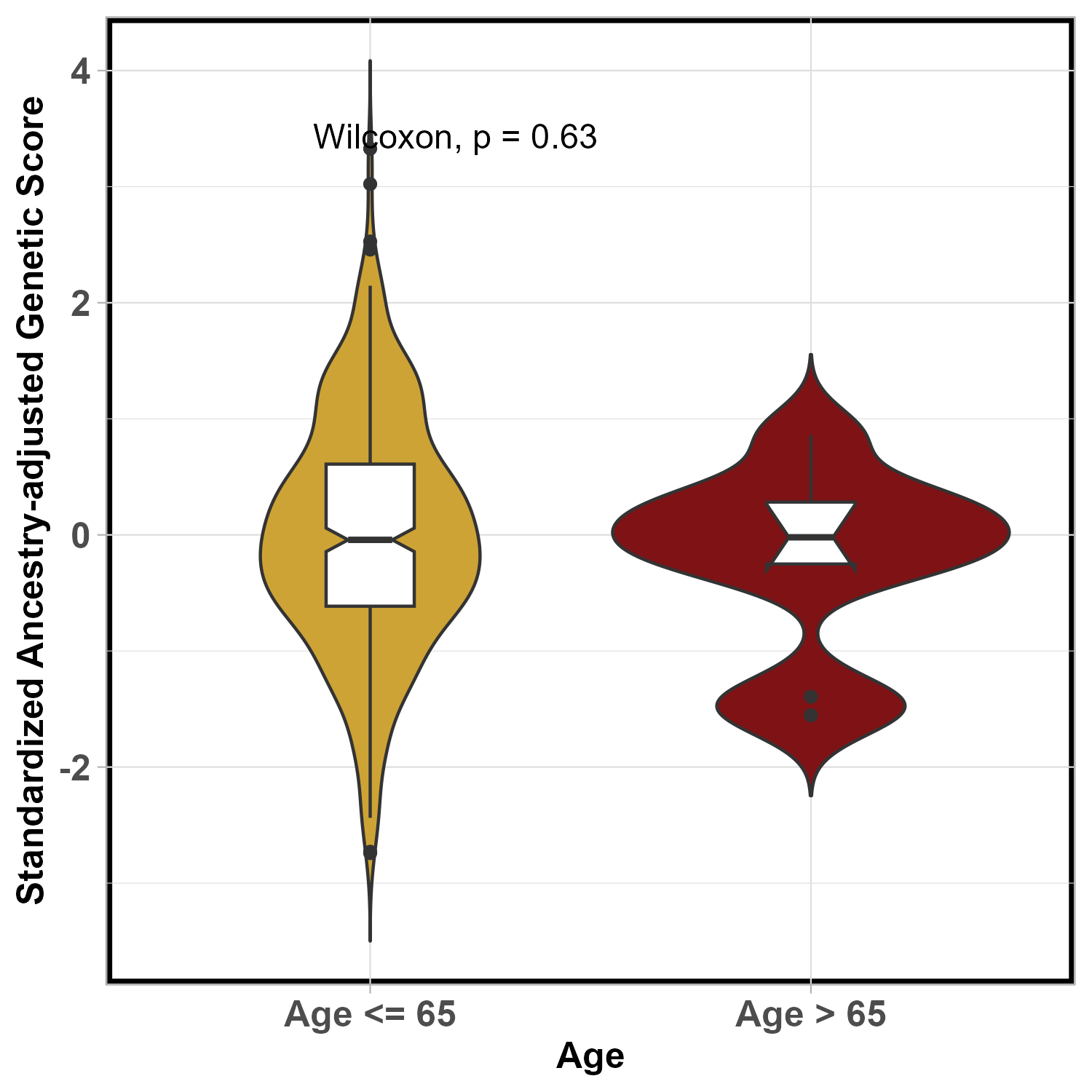

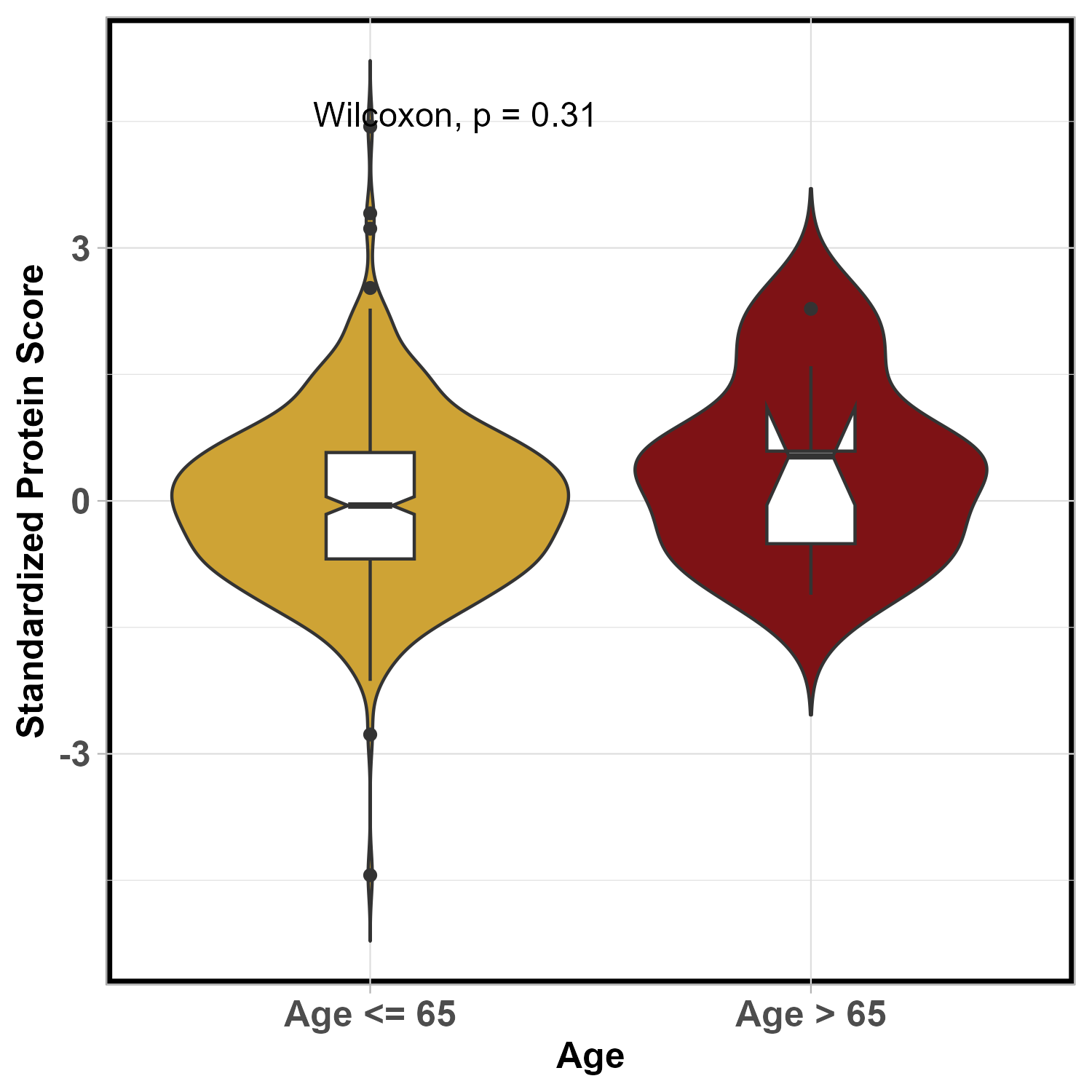

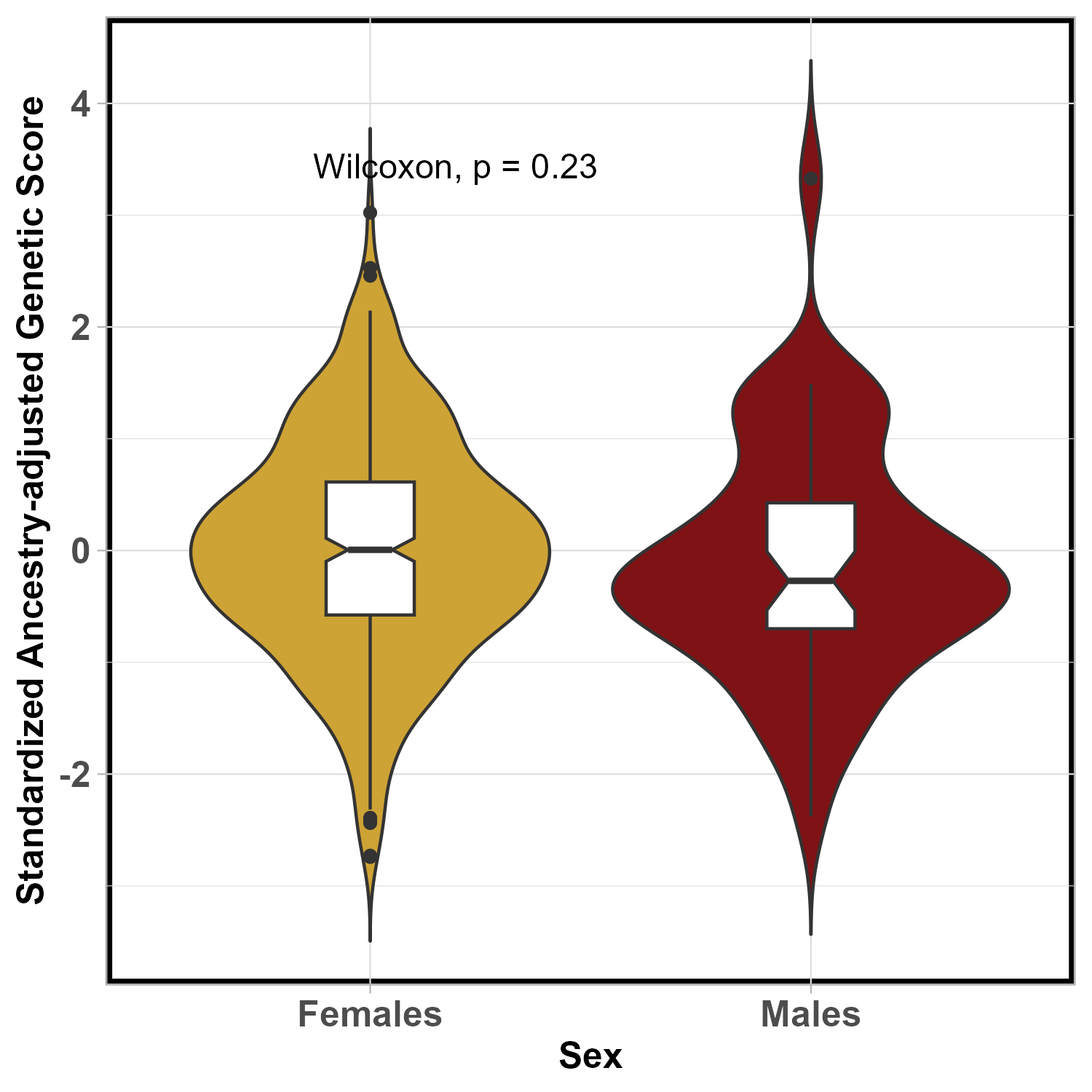

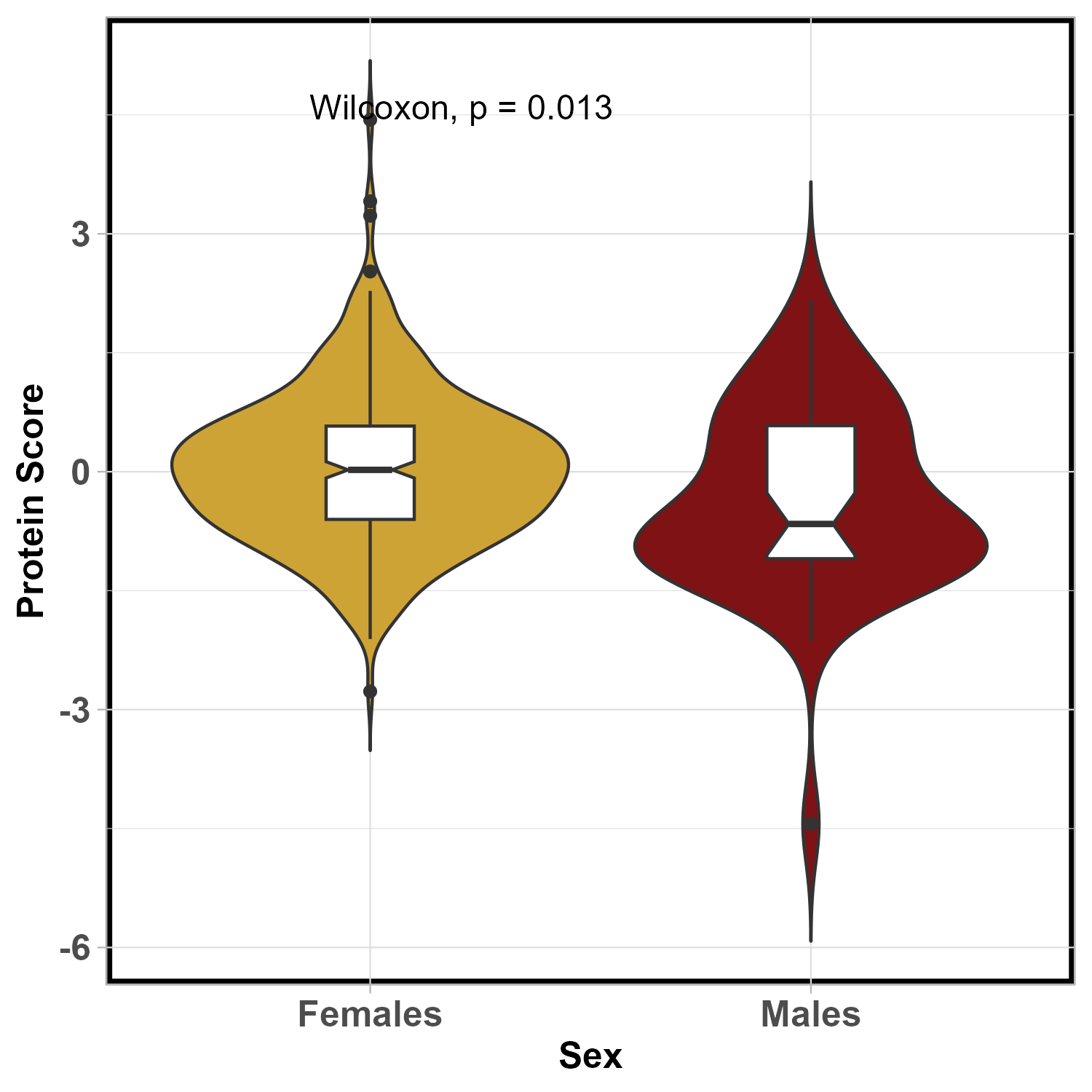

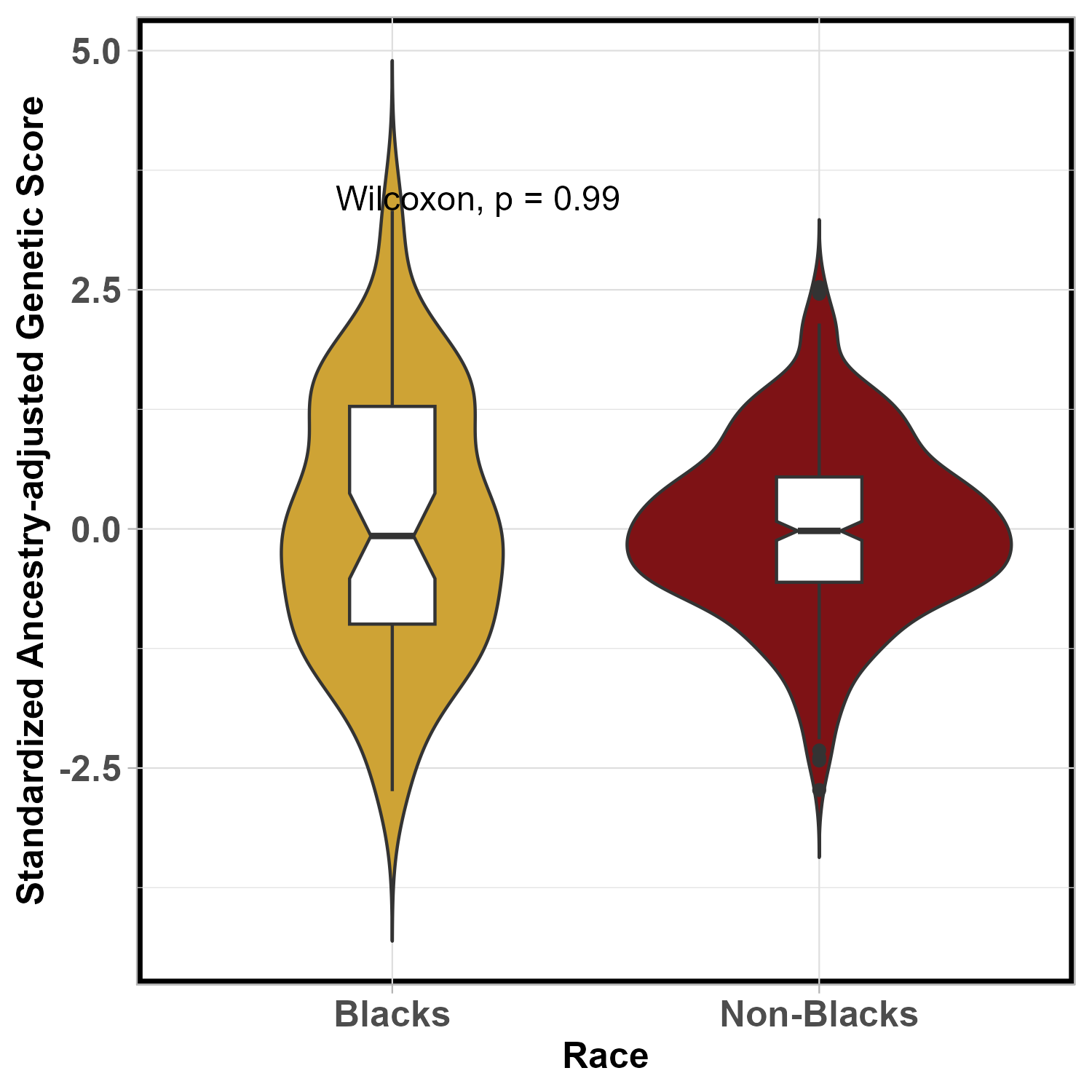

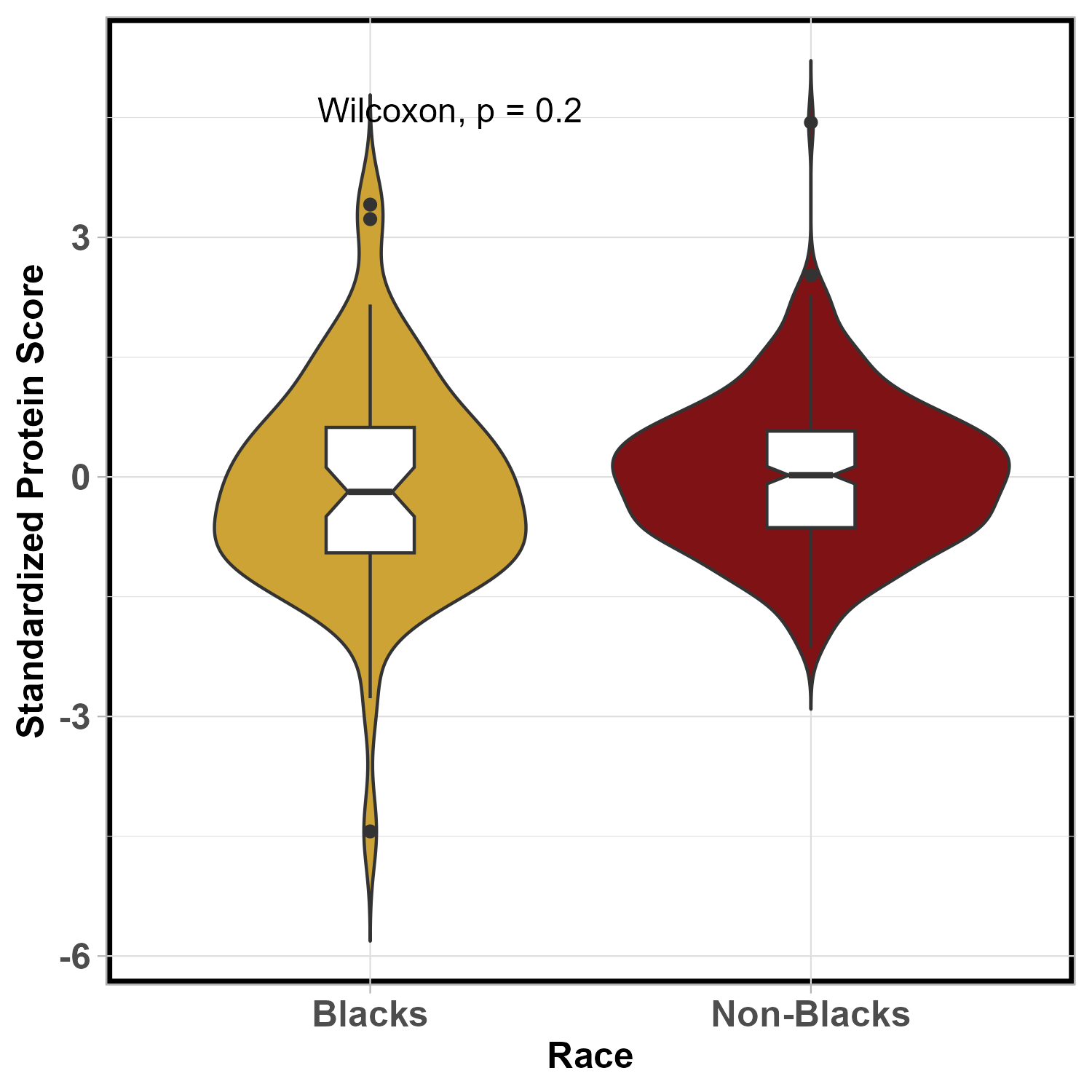
